# Decoding Treatment Choice: Genetic and Phenotypic Analyses of Long-term Antidepressant Acceptability

**DOI:** 10.1101/2025.08.19.25333784

**Authors:** Alicia Walker, Brittany L. Mitchell, Tian Lin, Jacob J. Crouse, Clara Albinana, Chloe X. Yap, Mary Ellen Lynall, Penelope A. Lind, Andrea Cipriani, Enda M. Byrne, Sarah E Medland, Nicholas G. Martin, Maxime Taquet, Ian B. Hickie, Naomi R. Wray

## Abstract

**Importance:** Antidepressant treatment remains largely trial-and-error, with approximately one-third of patients with major depressive disorder (MDD) reporting inefficacy of first-line medications. Identifying predictors of treatment acceptability is needed to improve prescribing precision and patient outcomes.

**Objective:** To identify phenotypic and genetic signatures associated with antidepressant acceptability and treatment complexity.

**Design, Setting, and Participants:** Retrospective cohort study of Australian Genetics of Depression Study participants dispensing ≥1 prescription of the 10 most commonly used antidepressants, based on linked 4.5-year (2013–2017) prescription data.

**Exposures:** Treatment complexity was assessed as number of antidepressant classes dispensed. Antidepressant acceptability was defined as sustained single-antidepressant use (≥360 cumulative days). Participants with genotyping data were classified into mutually exclusive groups based on sustained single-medication use.

**Main Outcomes and Measures:** Associations with 40 phenotypes and polygenic scores (PGS) for 15 traits. Genome-wide association studies (GWAS) were conducted for SSRI and SSRI/SNRI antidepressant acceptability.

**Results:** Of 13,763 participants with ≥1 antidepressant prescription, 9,844 had genotyping data (mean [SD] age 44.5 [15.0] years; 26% male; 89% lifetime MDD). Treatment complexity was significantly associated with 27 of 40 phenotypes (e.g., smoking, recurrent MDD, suicidal ideation, chronic pain, BMI) and higher PGSs for psychiatric traits (MDD, ADHD, bipolar disorder, neuroticism; β = 0.028– 0.045 per PGS SD unit, p = 9.9 × 10^-6^ to 2.1 × 10^-10^). Sixty percent met criteria for an exclusive antidepressant acceptability group. These groups had distinct phenotypic profiles, including associations with BMI, suicidal ideation, and comorbidities. In particular, only SNRI users had higher BMI than SSRI users, but this association was explained by the BMI PGS, indicating genetic rather than treatment-induced differences. GWAS identified novel loci including an immune-related gene *LY9*, for which the G allele of rs6427545 was associated with reduced SSRI acceptability (frequency = 0.31; OR = 0.82; p = 2.8 × 10^-8^)

**Conclusions and Relevance:** Antidepressant acceptability and complex treatment patterns correspond to distinct phenotypic and genetic signatures. PGS demonstrate potential for enabling precision psychiatry, and immune-related pathways warrant therapeutic investigation.

**Key Points:** *Question:* Can phenotypic and genetic factors help identify individuals likely to show antidepressant-specific acceptability and treatment complexity, improving treatment precision in major depressive disorder (MDD)?

**Findings:** In this cohort study of 13,763 individuals with antidepressant prescriptions, treatment complexity was associated with 27 self-reported phenotypic traits and higher polygenic scores (PGS) for psychiatric conditions. Among 9,844 genotyped participants, 60% met criteria for single-antidepressant acceptability, with distinct phenotypic and genetic profiles. Notably, higher BMI among sustained SNRI users was explained by genetic predisposition. A genome-wide association study identified novel loci, including an immune-related gene (*LY9*), associated with reduced SSRI acceptability.

**Meaning:** Phenotypic and genetic factors, including PGS, are associated with both antidepressant acceptability and treatment complexity. These findings support the use of such markers to guide treatment selection and identify patients at risk for more complex courses, informing precision psychiatry and early intervention in MDD.

## Introduction

Major Depressive Disorder (MDD) has a lifetime risk of ∼16% worldwide^1^. While antidepressants are more effective than placebo^2^, treatment selection remains largely empirical (“trial-and-error”). Clinicians must balance efficacy against tolerability while tailoring decisions to patients’ clinical profiles^3^. In Australia, prescribing guidelines (RANZCP, 2020) recommend initially selecting one of 7 first-line antidepressants, each from a different class, then sequentially trialling others from this group before considering alternative strategies^3^. Despite these structured approaches, international response rates remain suboptimal^4–6^. This challenge is evident in the Australian Genetics of Depression Study (AGDS), where around one-third of users rated each common antidepressant as ineffective, and 42% had trialled three or more^6^. These findings highlight the need for better predictors of treatment response.

Antidepressant outcomes likely reflect complex interactions between drug mechanisms and patient-specific biological or clinical factors. Genome-wide association studies (GWAS) estimate that 13-25% of variance in antidepressant response^7,8^ and 8% in treatment-resistant depression (TRD)^9^, as measured by clinical or self-report tools, are attributable to common genetic variation. While few genome-wide significant variants have been identified, polygenic scores (PGS), which aggregate small genetic effects, offer potential to predict clinical outcomes^10,11^. For example, schizophrenia PGS has been associated with poorer lithium response in bipolar disorder (BIP)^12^, suggesting a role for PGS in biologically informed treatment selection.

Prior research has used clinical outcomes (e.g., remission and symptom improvement), or self-report scales^7,8,13–15^ to investigate antidepressant response, and electronic health records (EHRs) to predict TRD^16^. However, these approaches often miss real-world indicators of treatment response. Centralised prescription databases provide an objective, scalable means to assess medication use and proxy outcomes^17^. Most studies using such data approximate antidepressant response via switching or sustained use^17–20^, though TRD definitions vary across studies^9,19^. To date, no study has directly compared genetic and phenotypic profiles across individuals stabilised on different antidepressants.

In this study, AGDS data linked to 4.5 years of national prescription records were used to investigate phenotypic and genetic correlates of real-world antidepressant acceptability. Individuals were classified into subgroups based on sustained use (≥360 cumulative days) of a single antidepressant, with sustained use acting as a proxy for real-world acceptability encompassing both tolerability and efficacy, since the lack of either typically prompts discontinuation or switching to another treatment strategy. This approach assumes that individuals with long-term single-medication use represent more biologically homogeneous subsets of MDD, potentially enabling identification of treatment-specific markers. In parallel, we explore proxies for difficult-to-treat depression (DTD)^21,22^, operationalised as antidepressant class diversity and total dispensed days to reflect treatment complexity and intensity. Overall, this represents the first use of AGDS data to assess antidepressant treatment outcomes via dispensation records, extending beyond earlier self-report-based assessment^23–26^.

## Methods

### Study Population and Data Sources

The Australian Genetics of Depression Study (AGDS)^6,27^ is a national volunteer cohort study examining contributors to depression and treatment responses (see details in Byrne, et al.^6^). During initial data collection, between April 2017 and March 2018, 22,414 participants aged ≥18 completed online questionnaires assessing psychosocial factors, lifetime MDD (via the Composite International Diagnostic Interview Short Form [CIDI]), and responses to the 10 most commonly prescribed antidepressants in Australia. Of these, 19,291 (88.9%) met criteria for lifetime MDD, with 724 (3.2%) having missing data. Participants provided saliva samples for genetic analysis using the Illumina Global Screening Array, with 14,501 individuals of genetically inferred European ancestry passing quality control^28^. Additionally, 14,471 AGDS participants consented to linkage with Australian Pharmaceutical Benefits Scheme (PBS) data through Services Australia, providing prescription data from July 1, 2013, to December 31, 2017. PBS data access was approved by Services Australia EREC committee (MI3967). Ethical approval for this study was obtained from the QIMR Berghofer Ethics committee (P2118).

### Study Medications and Cumulative Prescription Duration Estimation

To ensure broad cohort representation and robust subgroup analyses, we focused on the 10 most commonly prescribed antidepressants in Australia (under the N06A ATC Code) spanning four common classes (**Supplementary material S1**): (1) selective serotonin inhibitors (SSRIs): citalopram, escitalopram, paroxetine, sertraline, fluoxetine; (2) serotonin-norepinephrine reuptake inhibitors (SNRIs): desvenlafaxine, duloxetine, venlafaxine; (3) tricyclics (TCA)s: amitriptyline; and (4) tetracyclics (TeCAs): mirtazapine. This approach retained 94.5% of PBS-linked participants (N=13,673), ensuring representative coverage of antidepressant use in the cohort.

Due to PBS dosage reporting complexity, we initially assumed each prescription covered 30 days, based on typical Australian prescribing patterns. We then refined this estimate by using the lesser of 30 days or the interval until the next dispense of the same medication. This method helps prevent overestimation of treatment duration due to early refills or overlapping prescriptions used to achieve higher doses. Our approach emphasizes treatment duration rather than dosage intensity. Cumulative duration was calculated as the sum of individual prescription durations at the drug level, allowing for natural treatment gaps. Prescription episodes were defined as continuous medication dispensing periods, with new episodes beginning when gaps exceeded 90 days.

### Sustained Use Threshold as a Proxy for Antidepressant Acceptability

We defined treatment acceptability as sustained cumulative use (≥360 days) of a single antidepressant. This threshold was informed by RANZCP and NICE guidelines, which recommend ≥ 6 months of maintenance therapy to prevent relapse following initial treatment^3,29^. We selected 360 days (∼12 months) to conservatively capture both the acute treatment phase and the recommended maintenance period, representing a complete treatment cycle. A secondary ≥600-day threshold was used in sensitivity analyses to capture extended treatment, as recommended in recurrent depression^3,29^. While this method does not directly measure adherence, it reflects real-world prescribing behaviour. For instance, cases of non-continuous but recurrent use may still meet the cumulative threshold (see **Supp.** Fig 1). Importantly, this threshold is more stringent than common stable response cut-offs (≥180 days)^30^, and clinical trials often define acceptability by discontinuation rates after shorter periods (e.g., 8 weeks)^2,31^. Using a higher threshold helps generate more homogenous groups by reducing the proportion of individuals who discontinue treatment late due to sustained side-effects.

### Treatment Group Classification

We focused on 9,844 genotyped participants with European ancestry and ≥1 antidepressant prescription, and assigned to mutually exclusive treatment subgroups (**Supp Fig. 1**). These subgroups were defined based on treatment patterns. While participants to the AGDS study were recruited based on having depression, some of them self-reported a diagnosis of bipolar disorder (BIP). We did not exclude them from the analysis to keep the sample representative. However, we kept them in separate subgroups to account for diagnostic uncertainty and avoid bias from pseudo-treatment complexity. We further separated this BIP subgroup into those on and those not on Lithium as the former more clearly indicates a diagnosis of BIP and less diagnostic uncertainty.

**Figure 1.**
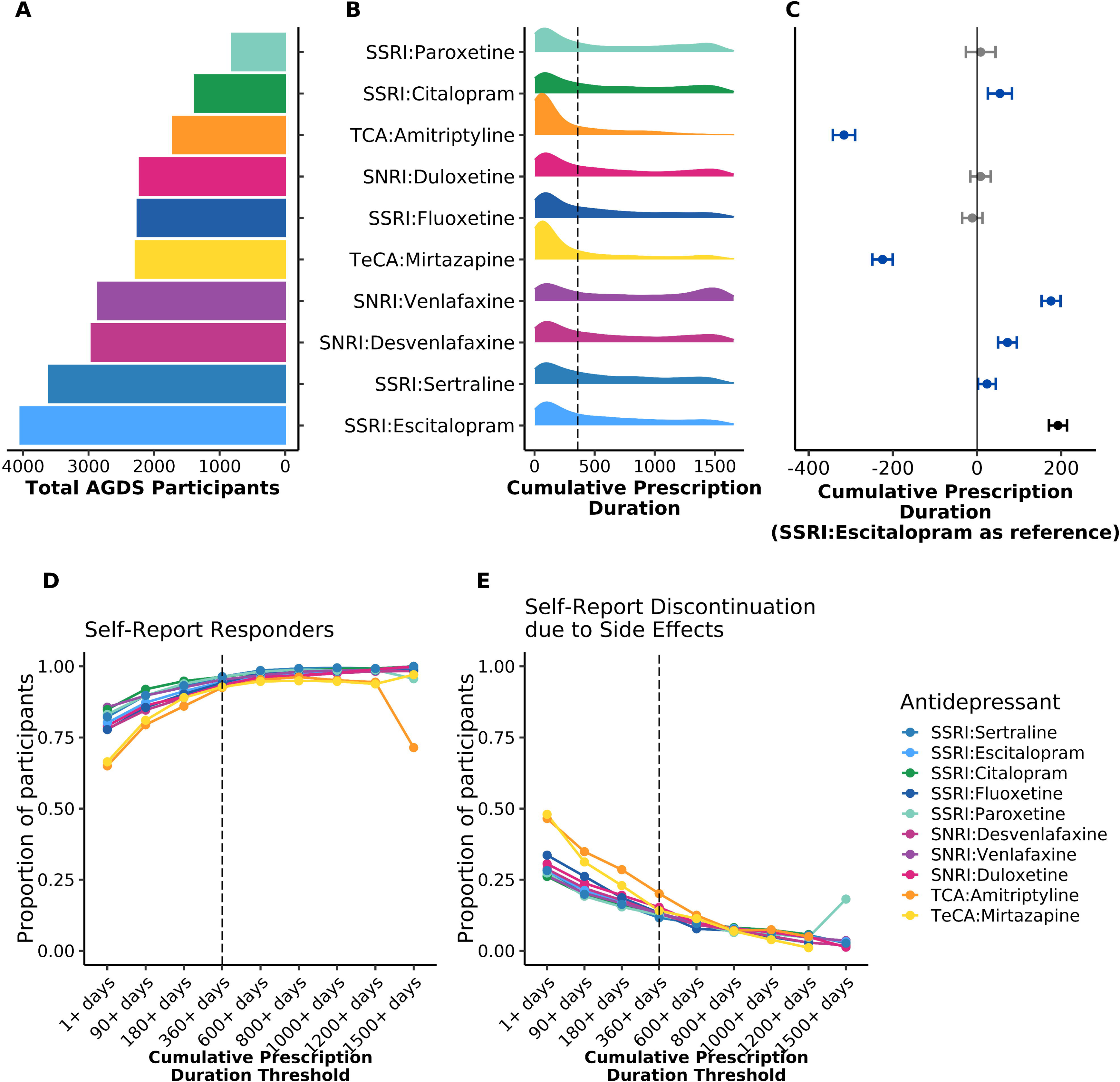
Antidepressant use and outcomes of the Australian Genetics of Depression Study (AGDS). Data are shown for 13,763 AGDS participants who had ≥1 recorded dispense for one of the 10 most commonly prescribed antidepressants (ADs) in Australia, based on linked Pharmaceutical Benefits Scheme (PBS) records from July 2013 to December 2018. The 10 most commonly prescribed ADs are summarized across five panels: (A) Number of participants with ≥1 recorded dispense for each AD; (B) Distribution of cumulative prescription duration per AD. The dashed line indicates the primary minimum threshold used to define AD acceptability; (C) Cumulative prescription duration relative to the reference AD, SSRI-escitalopram, estimated using linear regression adjusted for age and sex, across 13,693 participants. The marginal mean for escitalopram (reference AD) is shown in black (representing the baseline value); ADs with a statistically significant difference are shown in blue; non-significant differences are shown in grey; (D) Proportion of participants self-reporting a positive response to the dispensed AD, stratified by increasing thresholds of cumulative prescription duration (range: ≥1 to ≥1500 days). The dashed line indicates the acceptability threshold. Responses of “Moderately well” or “Very well” were classified as responders; “Not at all” as non-responders. (E) Proportion of participants self-reporting discontinuation due to any side effects, also stratified by cumulative prescription duration (range: ≥1 to ≥1500 days), with the same dashed line indicating the acceptability threshold.

This resulted in 5 subgroups (1) ‘Sustained single-antidepressant use’ without self-reported BIP; (2) ‘Combination’: sustained use of multiple antidepressants without self-reported BIP; (3) ‘Various’: other dispensing patterns without self-reported BIP; (4) ‘BIP with sustained lithium treatment’ (BIP+L: dispensed >3 lithium prescriptions over the 4.5-year period of pharmaceutical data); (5) ‘BIP without sustained lithium treatment’ (BIP-L: dispensed ≤3 lithium prescriptions). The first group was further stratified by specific antidepressant class, based on sustained use of ≥1 antidepressant within the same class. Those with sustained use across multiple classes were allocated to the ‘Various’ group.

### Statistical Analysis

The statistical analyses conducted in this study are detailed in the supplement. Briefly for the main analyses, linear and logistic regression models tested associations in two sets of analyses: (1) treatment complexity metrics as outcomes predicted by PGS for 15 traits and 40 self-reported phenotypes, with age and sex as covariates, and (2) PGS and phenotypes as outcomes predicted by antidepressant acceptability group assignment, with age and sex as covariates for self-report outcomes only. Each predictor was analysed separately against the outcome. In the second analysis, each group was systematically used as the reference but results are reported using SSRI/sertraline as reference groups.

## Results

### The Antidepressant Landscape of the Australian Genetics of Depression Study

Among the 10 most frequently dispensed antidepressants (13,673 AGDS participants), SSRIs and SNRIs were the most common, while TCAs and TeCAs had shorter cumulative treatment durations, likely reflecting their limited, adjunctive clinical use (**Supplementary Table 2, Fig. 1A–C**, class-level results in **Supplementary Fig. 2**). Validation analyses confirmed the ≥360 days cumulative prescription duration threshold as a good proxy for antidepressant acceptability: self-reported treatment response increased, and side effect burden decreased, steadily up to ∼360–600 days, then plateaued (**Fig. 1D–E**). Individuals with ≥360 days of treatment showed similar (stable) rates of recurrent depressive and prescription episodes compared to those with ≥180 or 600 days (**Supplementary Fig. 3**).

### Genetic and Phenotypic Factors Associated with Treatment Complexity

Half of 40 tested self-reported phenotypic characteristics were significantly associated with total antidepressant dispense after Bonferroni correction, with an even larger subset (68%) associated with medication diversity (**Supplementary Fig. 4, Supplementary Table 3**). Traits fell into three categories (**Supplementary Material S2**): those associated with prolonged use but not diversity of antidepressant classes (e.g., T2D, back pain), those linked to diversity but not overall dispense (e.g., smoking, lower education), and those associated with both greater dispense and diversity (e.g., recurrent MDD, suicidal ideation, chronic pain, BMI).

Next, we tested PGS associations between 15 traits and the sustained treatment groups (**Fig. 2, Supplementary Table 4**). PGS for major depression (MD) was not associated with total antidepressant dispense (we note that MD PGS is strongly associated with MDD status in AGDS compared to controls (OR=1.75, SE=0.017, p=7.34x10^-226^)^32^). However, it was significantly associated with increased class diversity (β=0.045 per SD PGS units, SE=0.0070, p=2.1x10^-10^).

**Figure 2.**
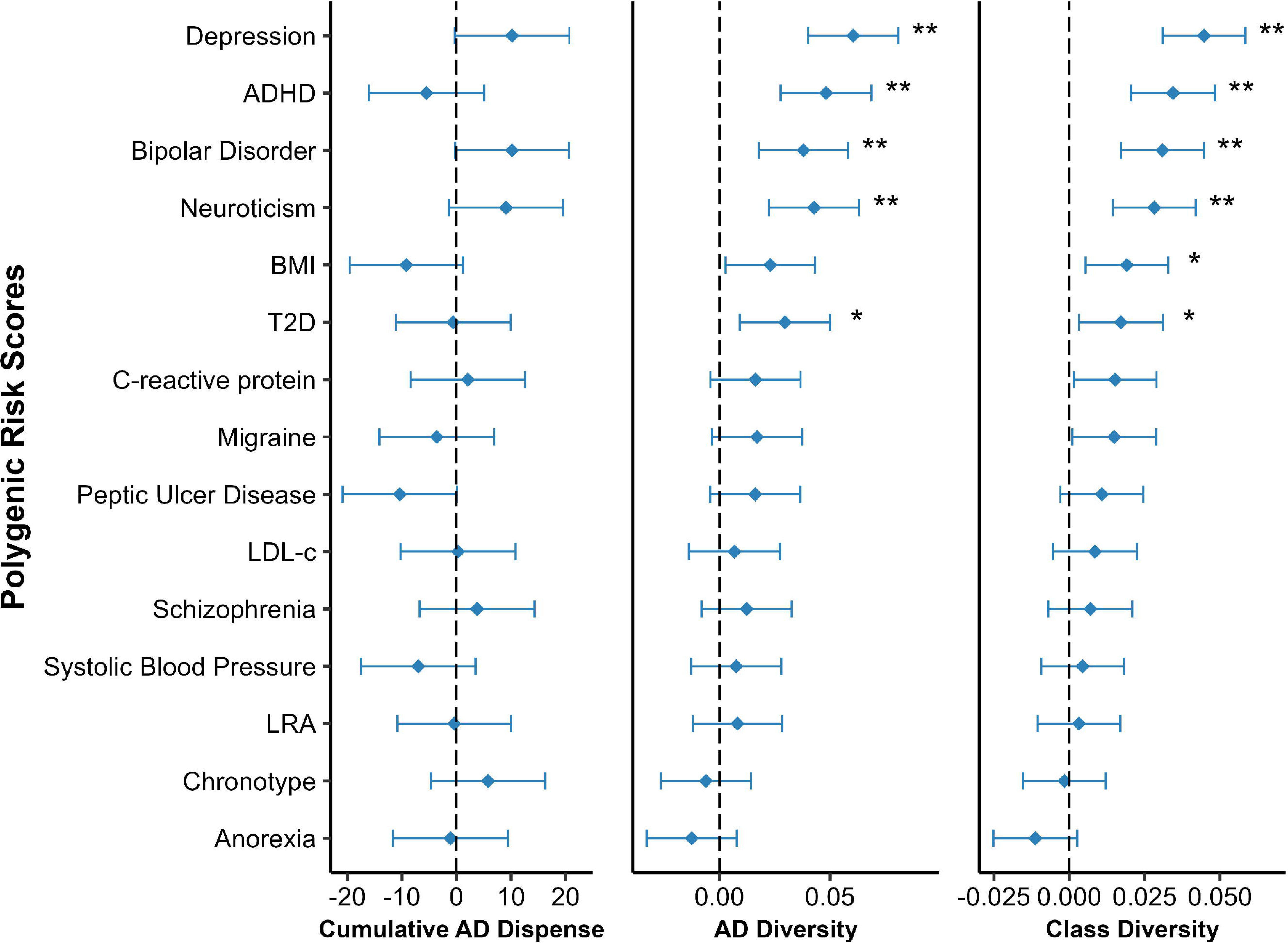
Associations between 15 polygenic scores and 3 antidepressant use metrics – cumulative antidepressant dispense, medication diversity, and class diversity - among 9,844 AGDS participants with valid genotyping data. (1) Left panel: Cumulative all antidepressant prescription duration (days); (2) Middle panel: antidepressant diversity (range: 1–10); (3) Right panel: Antidepressant class diversity (range: 1–4). All PGS associations are reported in standard deviation (SD) units, standardized across 14,603 AGDS participants of genetically inferred European ancestry. Associations were adjusted for age and sex. Statistical significance was declared at pL<L0.05 after false discovery rate (FDR) correction (*) and Bonferroni correction (**), applied separately within each DTD metric. See Supp. Table 4 for complete results, including the sensitivity analyses excluding individuals with self-report BIP.

Several psychiatric PGSs were also significantly associated with increased class diversity including ADHD (β=0.034, SE=0.0071, p=1.3x10^-6^), BIP (β=0.031, SE=0.0070, p=9.9 x 10^-6^), and neuroticism (β=0.28, SE=0.007, p=6.0x10^-5^). Sensitivity analyses excluding participants with self-reported BIP (n=1,066) confirmed these associations (**Supplementary Table 4**).

### Mutually Exclusive Antidepressant Acceptability Groups

Of genotyped participants, 5,891 (59.8%) demonstrated single antidepressant acceptability (**Supplementary Table 5**), with class-level allocations of 3,573 SSRI, 2,111 SNRI, 177 TeCA, and 151 TCA users (**Table 1, Supplementary Table 6**). Additionally, 846 participants had BIP-L, 220 had BIP+L, and 2,666 demonstrated various other patterns. Approximately 90% of participants in each group received a lifetime MDD diagnosis, with lower rates in TCA users (82%) and higher rates in bipolar groups (97%). Summaries under the more stringent threshold (600 days) to define antidepressant acceptability are in **Supplementary Tables 7-8**.

**Table 1.** Key AGDS participant summaries across antidepressant acceptability subgroups. Sustained use—used as a proxy for antidepressant acceptability—was defined as ≥360 cumulative days of dispensing. Treatment response was defined as a binary variable: participants reporting “Moderately well” or “Very well” were classified as responders, and those reporting “Not at all” as non-responders. All summaries are based on complete cases, with missing values (NAs) excluded from the denominator to avoid underestimating prevalence. See Supp. Tables 5-8 for complete results.

|  | *SSRI* | *SNRI* | *TCA* | *TeCA* | *BIP+L* | *BIP-L* | *Various* |
| --- | --- | --- | --- | --- | --- | --- | --- |
| *N* | 3573 | 2211 | 151 | 177 | 220 | 846 | 2666 |
| *Lifetime MDD (%)* | 88 | 91 | 82 | 90 | 97 | 97 | 91 |
| *Total Lifetime MDD score (median [SD])* | 8 (1.2) | 8 (1.1) | 8 (1.2) | 8 (1.1) | 9 (0.7) | 9 (0.8) | 8 (1.1) |
| *Age (yrs, mean [SD])* | 45 (15) | 46 (15) | 54 (14) | 51 (15) | 44 (13) | 44 (14) | 42 (16) |
| *Males (%)* | 23 | 25 | 26 | 48 | 30 | 28 | 29 |
| *Age of MDD onset (yrs, mean [SD])* | 23 (11) | 23 (12) | 28 (14) | 26 (14) | 21 (11) | 20 (9.9) | 23 (12) |
| *Number of MDD episodes (median [SD])* | 6 (4.4) | 7 (4.4) | 6 (4.6) | 10 (4.6) | 13 (3.8) | 13 (3.8) | 6 (4.5) |
| *Suicidal thoughts (%)* | 64 | 71 | 55 | 73 | 88 | 85 | 67 |
| *Self-harm (%)* | 36 | 42 | 33 | 35 | 57 | 64 | 43 |
| *BMI (mean [SD])* | 29 (6.8) | 29 (6.9) | 29 (7.1) | 29 (6.3) | 30 (7.0) | 30 (6.9) | 28 (7.0) |
| *T2D (%)* | 3.8 | 4.3 | 11 | 7.3 | 2.7 | 4.6 | 2.9 |
| *Chronic pain (%)* | 10 | 13 | 34 | 14 | 16 | 16 | 12 |
| *Migraines (%)* | 68 | 69 | 79 | 62 | 69 | 75 | 67 |
| *Education level (yrs, median [SD])* | 6 (1.1) | 6 (1.1) | 5 (1.2) | 5 (1.2) | 6 (1.1) | 6 (1.2) | 6 (1.1) |
| *Physical health level (median [SD])* | 3 (3.3) | 3 (3.2) | 3 (3) | 3 (3.2) | 3 (3.1) | 3 (3.1) | 3 (3.2) |
| *Regular smoker (%)* | 72 | 73 | 82 | 79 | 77 | 80 | 70 |
| *Drinks per 3 months (mean [SD])* | 14 (23) | 13 (24) | 9.9 (21) | 14 (26) | 11 (21) | 14 (25) | 12 (22) |
| *Self-report response to specific AD (%)* | 98 | 97 | 94 | N/A | N/A | N/A | N/A |
| *Self-report response to any AD (%)* | 100 | 100 | 100 | N/A | 100 | 100 | 100 |
| *Self-report discontinuation to AD (%)* | 9.3 | 9.6 | 17.3 | 12.1 | N/A | N/A | N/A |
| *Self-report discontinuation to any AD (%)* | 100 | 100 | 100 | N/A | N/A | N/A | N/A |

### Participant Phenotypes Differentially Associate with Choice of Antidepressant Medication

Dispensing patterns and phenotypic factors showed distinct class-specific associations (**Fig. 3, Supplementary Material S3-4; Supplementary Fig. 5-9, Supplementary Tables 9-12**). Key patterns (relative to SSRI users) included: SNRI users showed higher BMI and more severe psychiatric comorbidity; TCA users were older with prominent somatic conditions; TeCA users had elevated suicidal ideation and male overrepresentation; and BIP groups showed distinct psychiatric profiles based on lithium treatment status.

**Figure 3.**
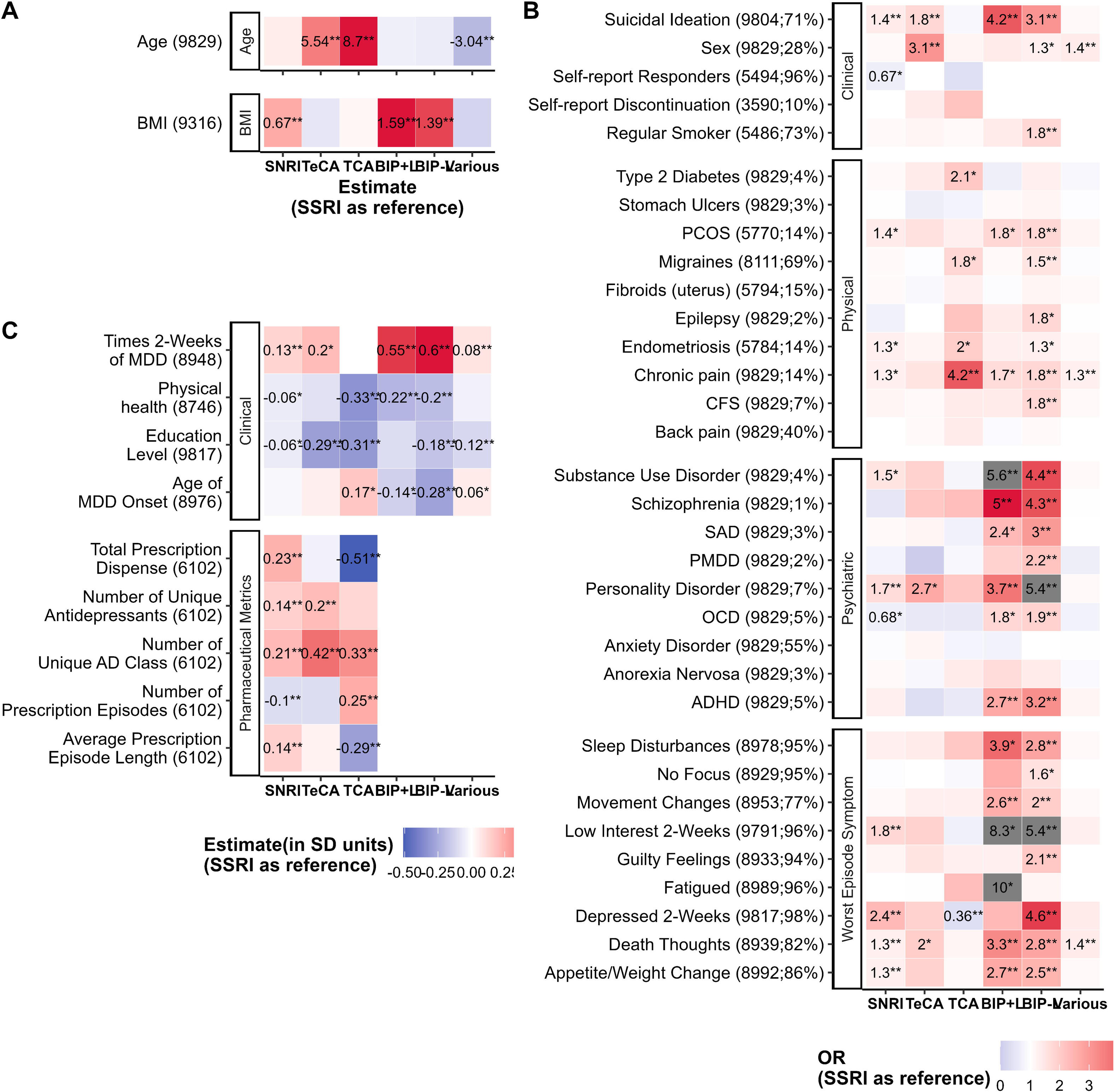
Heatmap of antidepressant dispense patterns and self-reported characteristics associated with antidepressant acceptability groups. Dependent variables associated across treatment groups included: (A) unscaled age and BMI; (B) scaled binary outcomes spanning phenotypic, physical, and psychiatric traits, as well as symptoms during the worst MDD episode. The y-axis reports the number of AGDS participants with complete data for age, sex and the binary trait, as well as % endorsing the disorder/trait; and (C) scaled quantitative outcomes related to pharmaceutical metrics and phenotypic features. Grey boxes indicate odds ratios (OR) > 5. All dependent variables (except age and BMI) were standardized (mean = 0, SD = 1) across the entire AGDS cohort with ≥1 antidepressant (AD) dispense (N = 13,763). All models included age and sex as covariates, except when these variables were the outcomes of interest. The SSRI group served as the reference category. Statistical significance was determined at p < 0.05 after correction for multiple testing: false discovery rate (FDR; *) and Bonferroni correction (**), applied separately within each AD class. For interpretability: higher physical health scores indicate better health; higher education scores indicate greater educational attainment; and “female” was the reference group for sex. See Supp. Tables 9–12 for complete results.

### Polygenic Risk Scores Differentially Associate with Choice of Antidepressant Medication

Next, we assessed associations between antidepressant treatment groups and 15 PGS for psychiatric, cardiometabolic, and circadian traits (**Fig. 4A, Supplementary Fig. 10, Supplementary Tables 13-14**). No antidepressant group differed significantly in MD PGS relative to SSRIs, consistent with MD GWAS cases comprising a mix of our antidepressant class groups. In contrast, BIP PGS was significantly higher in both lithium-treated (BIP+L: β = 0.280 per SD unit of the AGDS cohort, SE = 0.069, p = 5.6 × 10^-5^) and non-lithium-treated (BIP−L: β = 0.323, SE = 0.038, p = 3.5 × 10^-17^) subgroups compared to SSRIs. The BIP−L group showed broad psychiatric polygenic vulnerability, with elevated PGS for MDD, SCZ, and ADHD compared to the SSRI group after Bonferroni correction, while BIP+L showed no significant associations with these psychiatric PGS. Multinomial regression further indicated that BIP PGS was significantly associated with TeCA use over SSRIs in the PGS-only model (β = 1.21, p = 0.015) (**Supplementary Table 17, Supplementary Fig. 12**). Beyond psychiatric traits, the BIP-L group also had significantly higher PGS for peptic ulcer disease (PUD) after Bonferroni correction (β = 0.099, SE = 0.038, p =9.7 x 10^-3^), while lower PUD PGS was nominally associated with SNRI over SSRI use in the combined PGS and self-report multinomial regression model (**Supplementary Table 17, Supplementary Fig. 12**).

**Figure 4.**
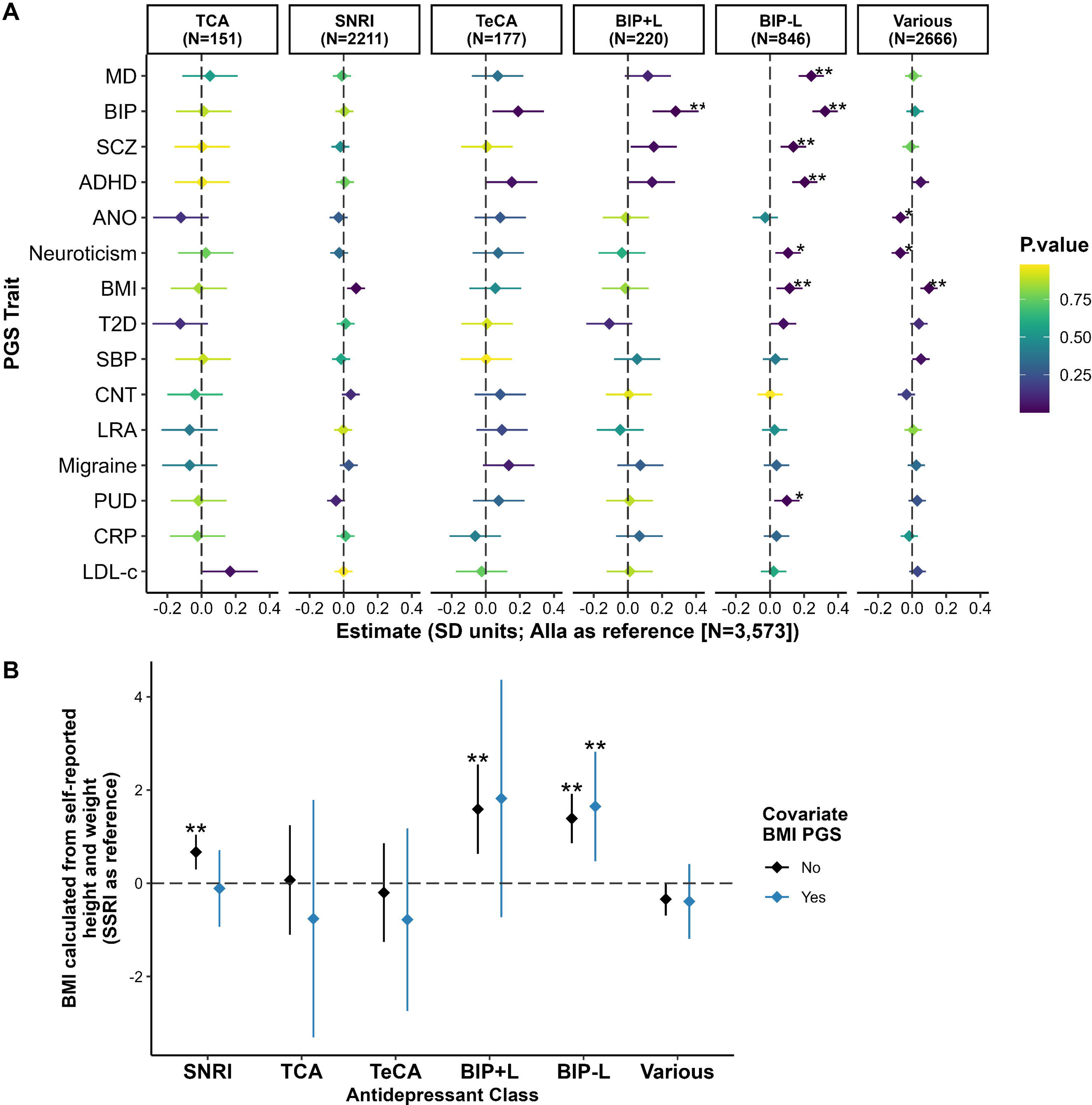
Polygenic and BMI profiles across antidepressant acceptability treatment groups. (A) Associations between 15 PGS and treatment groups were tested. All PGS were standardized (mean = 0, SD = 1) across the 14,603 AGDS participants of genetically inferred European ancestry. (B) Associations between BMI, calculated from self-reported height and weight, and treatment groups. Black estimates represent model results adjusted for age and sex; blue estimates represent results additionally adjusted for BMI PGS (also standardized across the same AGDS cohort). Statistical significance was declared at p < 0.05 after false discovery rate (FDR; *) and Bonferroni correction (**), applied separately within each antidepressant (AD) class. See Supp. Tables 13-16 for complete results.

In cardiometabolic domains, the SNRI group had a nominally higher BMI PGS relative to SSRIs (β = 0.073, SE = 0.027, p = 7.7 x 10^-3^) (**Fig. 4A, Supplementary Table 13**), particularly in the duloxetine (β = 0.139, SE = 0.054, p = 0.010) and venlafaxine (β = 0.116, SE = 0.047, p = 0.015) subgroups (**Supplementary Fig. 10, Supplementary Table 14**). This finding was supported by the multinomial regression in the PGS-only model (**Supplementary Table 17, Supplementary Fig. 12**), where BMI PGS was nominally associated with SNRI use over SSRIs (β = 1.07, p = 7.7 × 10^-3^). Among sustained antidepressant-use groups, only two PGS associations remained significant after Bonferroni correction. Systolic blood pressure (SBP) PGS was significantly lower in the SSRI-escitalopram group compared to the SSRI-sertraline group (β=-0.126, SE=0.043, p=3.1 × 10^-3^), while at the stricter 600-day sustained use threshold, the BMI PGS was significantly higher in the SNRI-duloxetine group compared to the SSRI-sertraline group (β=0.18, SE=0.057, p=0.0016) (**Supplementary Fig. 10, Supplementary Table 14**).

Given that BMI and its PGS are specifically associated with treatment choices, we ran additional analyses to disentangle the roles of this trait and its genetic predisposition. Adjustment for BMI PGS fully attenuated the Bonferroni-significant BMI elevation in SNRI users from β = 0.67kg/m^2^ (SE= 0.19, p = 4.0 × 10^-4^) to β=-0.11kg/m^2^ (SE=0.42, p=0.80) (**Fig. 4B, Supplementary Table 15**), indicating genetically driven rather than treatment-induced weight profiles. Moreover, BMI PGS fully attenuated the observed Bonferroni-significant BMI elevation in duloxetine users (**Supplementary Table 16, Supplementary Fig. 11**). BMI elevations in the BIP+L group remained stable post-adjustment, suggesting lithium (or confounding factors) contributes to higher BMI^33^. In the BIP-L group, BMI effect size was higher after PGS adjustment, suggesting BMI is genetically associated with group membership.

### GWAS on SSRI and SNRI Sustained Use and Self-reported Efficacy

GWAS comparing individuals with SSRI or SNRI acceptability (n=5,554) to those without (n=2,270) showed no inflation (λ = 0.997) and significant SNP-based heritability (**Supplementary Fig. 13**). One independent genome-wide significant locus was identified with top SNP rs11672755 (T allele; frequency = 0.26) associated with greater acceptability (OR=1.27, p=5.7× 10^-9^) (**Supplementary Fig 14, Supplementary Table 18**). This SNP is located in intron 2 of *DPRX*, encoding a PRD-like homeodomain transcription factor that represses genes activated during human preimplantation development^34^. SSRI-only acceptability GWAS (λ = 0.999, n cases=3,423, n controls=4,561) had one genome-wide significant locus (**Supplementary Fig. 15**). The lead SNP, rs6427545, in intron 2 of *LY9* (*SLAMF3*/*CD229*) showed the G allele (frequency = 0.31) associated with reduced SSRI acceptability (OR = 0.82, p = 2.8 × 10^-8^) (**Supplementary Fig. 16, Supplementary Table 19**). *LY9* encodes a cell surface receptor involved in the activation of lymphoid cell (B, T and NK cells)^35^, and is highly expressed in lymphoid tissues (protein atlas)^36^. However, the regulatory architecture of rs6427545 is complex, functioning as both an expression QTL and splicing QTL (e/sQTL), with tissue-dependent effects on multiple genes including *LY9, CD224, TSTD1, ITLN2*, and *NECTIN4*^37^. In EBV-transformed lymphocytes, the G allele shows increased splicing efficiency for some *LY9* transcripts but decreased protein expression, making net regulatory effects unclear. SNPs at the *LY9* locus retained suggestive significance (lead SNP rs790624: OR = 0.81, p = 6.1 x 10^-8^) in the combined SSRI/SNRI GWAS (**Supplementary Table 19**). Neither SSRI/SNRI nor SSRI self-reported efficacy GWAS identified genome-wide significant associations (**Supplementary Tables 20–21, Supplementary Fig. 17–18**).

## Discussion

While prior research has focused on symptom-based and molecular-based sub-typing^28,38–42^, we propose a novel approach to MDD sub-typing grounded in real-world prescription data. By sub-typing MDD into mutually exclusive sustained single-drug use (≥360 cumulative days) subgroups, we provide a scalable and pragmatic proxy for antidepressant-specific acceptability. This framework aims to parse the extensive diagnostic^43^, genetic and phenotypic heterogeneity^44^ of MDD and inform future precision psychiatry efforts.

In summary, beyond drug-level differences, psychiatric PGSs—including MD, ADHD, BIP, and neuroticism—were associated with greater antidepressant class diversity, highlighting a genetic contribution to treatment complexity and DTD, consistent with prior findings linking ADHD and neuroticism PGS to treatment-resistance^9,45^. The strong association between the MD PGS and greater treatment complexity supports biological mediation of complexity, rather than diagnostic uncertainty or misdiagnosis. After excluding participants with self-reported BIP, the BIP PGS was still significantly associated with treatment complexity, supporting the notion that undiagnosed BIP may contribute to pseudo-TRD^46^. Corresponding self-reported traits like personality disorder (often related to neuroticism^47^) and ADHD were linked to greater class diversity, with all traits except ADHD also tied to longer treatment duration, suggesting overt psychiatric comorbidity may contribute more to prolonged treatment than genetic risk. Within the AGDS bipolar subgroups, BIP-L participants showed significantly higher polygenic burden for MD, SCZ, ADHD, and neuroticism than SSRI users. BIP+L users did not show this pattern. This aligns with literature showing associations between lithium non-response and elevated PGS for SCZ^12^, MDD^48^, and ADHD^49^, and supports the hypothesis that lithium responders may have a less complex psychiatric profile^50^. Although direct comparisons between BIP-L and BIP+L were not statistically significant, likely due to sample size differences. Importantly, psychiatric PGSs did not differ across sustained antidepressant-use groups by drug class, reinforcing their role as perhaps stronger trans-diagnostic biological indicators of treatment complexity rather than predictors of specific pharmacologic response.

Metabolic genetic variation also influenced treatment patterns. Individuals with higher BMI were more likely to be sustained users of SNRIs (particularly duloxetine), than SSRIs. These effects did not persist after adjusting for BMI PGS, suggesting genetic metabolic liability may drive SSRI non-acceptability and/or SNRI tolerability. In contrast, higher BMI in BIP+L users remained after adjustment, consistent with lithium’s known metabolic side effect^33^, while sustained users of TeCAs and TCAs showed no elevation in BMI PGS or self-report BMI. Duloxetine has been recommended in Australia as a first-line agent for neuropathic pain (including diabetic neuropathy)^51^, and speculatively, alternative agents (e.g., duloxetine) may better suit individuals with MDD and elevated metabolic PGSs. Broadly, BMI and T2D PGS were associated (under FDR correction) with greater treatment diversity, mirroring psychiatric PGSs patterns. These findings support emerging evidence that metabolic genetic liability contributes to DTD, including prior reports of association of TRD and obesity PGS^9^, as well as with the *FTO* loci^16^.

We also report the first genome-wide significant associations with antidepressant acceptability using prescription data as the sole outcome. This contrasts with our null findings using self-reported response measures and underscores the advantage of a more stringent, homogeneous phenotype. One of the significant loci implicates immune signalling, with rs6427545 located within *LY9*, a cell-intrinsic regulator of T cell activation^52^, while promoting Th1 polarization and IFN-γ production^53^. Transdiagnostic genetic risk for psychiatric disorders implicates T cell activation in pathogenesis^54^, and SSRIs have been shown to have both inhibitory and stimulatory effects on T cell proliferation, dependent on context^55^. Altered *LY9* function could promote a lymphoid-linked immune sub phenotype of depression, or *LY9* may modify antidepressant-induced changes in T cell function, leading to altered drug acceptability. This SNP is in strong linkage disequilibrium (r² = 0.97) with another variant linked to reduced rTMS response in TRD^56^, further supporting its role in antidepressant response phenotypes. Our acceptability GWAS did not replicate the four genome-wide significant variants from a recent SSRI/SNRI response GWAS meta-analyses (only rs1106260 reached nominal significance) (**Supplementary Table 22**)^26^. Larger sample sizes or orthogonal data are needed to draw strong conclusions about the statistical similarity or biological validity of group constructs. Overall, our study suggests immune dysregulation may underlie a biologically distinct subtype of depression marked by treatment acceptability. Future *in vitro* studies should examine how *LY9* variants affect T cell responses to antidepressants.

In our context, long-term acceptability refers to the sustained real-world use of a specific antidepressant, reflecting a balance of tolerability, adherence, and patient- or clinician-perceived benefit over time. Importantly, the sustained use groups had lower rates of self-reported discontinuation due to side-effects for their assigned antidepressant compared to any other antidepressants they had tried (9–17% vs 100%) but had similar rates of self-reported efficacy (**Table 1**). This suggests that sustained use reflects antidepressant-specific tolerability more than efficacy.

Limitations in our study include the 4.5-year prescription window and restriction to the top 10 antidepressants, which do not capture lifetime treatment trajectories. However, follow-up work shows high consistency in group assignments over time (Wray, unpublished). Prescribing also reflects clinician judgment, not just biology, and may evolve with changing guidelines, perhaps introducing forms of selection bias. Replication in large, genotyped cohorts like Lifelines^57^, UK Biobank^58^, and iPSYCH^59^ will be essential to validate our findings and distinguish biological from clinical influences on antidepressant acceptability. Third, PBS records reflect dispensing, not adherence, and do not measure symptom change or quality of life to capture therapeutic benefit. Additionally, reasons for discontinuation (side effects vs. lack of efficacy) cannot be determined from dispensing records alone. Fourth, online recruitment and self-report data may introduce bias, and non-pharmacologic interventions (e.g., psychotherapy) were not accounted for. Finally, small sample sizes in TeCA, TCA, and BIP+L groups limited power, and PGS performance varies across traits due to GWAS characteristics (we benchmarked power via number of genome-wide hits, sample size, and SNP-based heritability [**Supplementary Table 1]**).

In conclusion, our findings support real-world sustained antidepressant-use as a phenotypically and genetically informative approach to depression stratification. Observed associations, such as higher suicidality in TeCA users, somatic symptoms in TCA users, and elevated BIP PGS in BIP-flagged groups, highlight the validity of our classifications. The relationship between SNRI treatment acceptability and BMI PGS, along with increased polygenic risk for common disorders in complex treatment patterns, suggests that PGS may inform treatment selection beyond traditional diagnostic categories, and act as objective markers for patients with DTD who may benefit from early intensive intervention. While replication of the GWAS results is needed, the data support a role for immune modulation in antidepressant response. These findings highlight the therapeutic potential of immune-related genetic markers in precision psychiatry.

## Data Availability

Data used in this analysis and described in this article are available, upon reasonable request, to all interested researchers through collaboration. Please contact NGM ( Nick. Martin@ qimrberghofer. edu. au), noting that access to the PBS data is not available to external researchers.

## Code Availability

All code to run the analysis can be found on the GitHub Page: https://github.com/walkeralicia/Antidepressant_Acceptability

## Data Availability

Data used in this analysis and described in this article are available, upon reasonable request, to all
interested researchers through collaboration. Please contact NGM ( Nick. Martin@
qimrberghofer. edu. au), noting that access to the PBS data is not available to external researchers.

## Acknowledgements and Competing Interests

We are indebted to all of the AGDS participants for giving their time to contribute to this study. We thank all the people who helped in the conception and implementation of the study. The AGDS was primarily funded by National Health and Medical Research Council (NHMRC) of Australia grant 1086683. The record linkage to the PBS data was funded by NHMRC Investigator grant 1172917. This study was further supported by NHMRC grants 1173790, 1073898 (NRW) 1172917 (SEM) 2017176 (BLM) 1172990 (NGM), 2008197 (JJC). AW and NRW are funded by the Michael Davys Trust at the University of Oxford. NRW also acknowledges funding for the AMBER: Antidepressant Medications: Biology, Exposure & Response study from the Wellcome Trust.

Professor Hickie is a Professor of Psychiatry and the Co-Director of Health and Policy, Brain and Mind Centre, University of Sydney. He has led major public health and health service development in Australia, particularly focusing on early intervention for young people with depression, suicidal thoughts and behaviours and complex mood disorders. He is active in the development through codesign, implementation and continuous evaluation of new health information and personal monitoring technologies to drive highly-personalised and measurement-based care. He holds a 3.2% equity share in Innowell Pty Ltd that is focused on digital transformation of mental health services.

## Notes

### Author Declarations

The External Request Evaluation Committee (EREC) of Services Australia gave ethical approval of this work. The Ethics committee of QIMR Berghofer gave ethical approval of this work.

