## Supplementary Text for "Decoding Treatment Choice: Genetic and Phenotypic Analyses of Long-term Antidepressant Acceptability"

Supplementary Material Table of Contents

### Methods

#### Polygenic Score Calculation

As comorbidities and side-effects influence clinician antidepressant selection, we calculated polygenic scores (PGS) to assess genetic liability for 15 traits linked to depression or BIP, or their treatment response (**Supp. Table 1**). PGS were estimated for major depression (MD; defined as MD because the GWAS combined MDD cases with self-report depression), and common comorbid psychiatric and developmental conditions, including attention deficit hyperactivity disorder (ADHD), anorexia nervosa (ANO), bipolar disorder (BIP), schizophrenia (SCZ), obsessive-compulsive disorder (OCD), and neuroticism. Given the frequent occurrence of metabolic syndrome, autoimmune conditions, and migraines in MDD and its treatment, we also calculated PGS for body mass index (BMI), type 2 diabetes (T2D), systolic blood pressure (SBP), migraines, C-reactive protein (CRP) levels, and low-density lipoprotein cholesterol (LDL-c). Additionally, we included PGS for chronotype, as it is associated with MDD^1^, and low relative amplitude (LRA) due to the association between circadian rhythms and lithium response in BIP^2,3^. Based on evidence suggesting a causal relationship between MDD and peptic ulcer disease (PUD)^4^, we also calculated a PGS for PUD.

PGS were computed using SBayesRC^5^ with GCTB software^6^, employing a Bayesian multiple regression framework that integrates functional annotations to optimize SNP weights. Input included GWAS summary statistics and an LD matrix calculated from 20,000 randomly selected UK Biobank genetically inferred European individuals covering ~7.4 million SNPs. Using PLINK2^7^, we multiplied joint effect estimates by imputed best guess genotypes and summed values to create PGS, standardized (mean=0, SD=1) across all 14,603 genetically inferred European ancestry participants.

#### Antidepressant Treatment Complexity

Before group classification, we examined associations between 40 self-reported traits and 15 PGS with three treatment complexity metrics, reflecting components of DTD, across all 13,673 participants with ≥1 antidepressant prescription for at least one of the 10 study medications. Considering only prescriptions for these 10 antidepressants, these complexity metrics included: (1) cumulative prescription duration (days) for all 10 antidepressants combined; (2) medication diversity (unique antidepressant count [range: 1-10]) and (3) class diversity (unique antidepressant class count [range: 1-4]). Linear regression models included age at baseline and sex as covariates. Nominal statistical significance was defined as p < 0.05. Given correlation between tests, results are reported significant based on an FDR and more stringently after Bonferroni correction, applied separately across each complexity metric. Sensitivity analyses excluded 1,066 participants with self-reported BIP to account for diagnostic uncertainty, resulting in a final analytic sample of 12,607.

#### Treatment Group Comparisons

Linear and logistic regression models were used to test differences across treatment groups at drug and class levels. Effect sizes are reported as beta coefficients (+/- SE) for linear models and odds ratios (with 95% CI) for logistic models. All self-report quantitative variables except age and BMI were standardised (mean = 0, SD =1) across the entire AGDS cohort with ≥1 antidepressant dispensed (N = 13,763). Models were specified as: PGS ~ Treatment_Group, and Outcome ~ Age + Sex + Treatment_Group. To obtain all pairwise comparisons between antidepressants (as a results resource in the supplementary material each drug was systematically used as the reference group in turn. However, for consistency in visualisations, we report associations from models using the SSRI group (for class-level comparisons) and the sertraline group (for drug-level comparisons) as the reference groups, given they are the most commonly prescribed antidepressants in Australia. Results are reported significant based on FDR and more stringently after Bonferroni correction, applied within each treatment group. For BMI analyses, additional models included BMI PGS adjustment to assess genetic independence.

Multinomial logistic regression models evaluated predictors of antidepressant class assignment (SSRI, SNRI, TeCA, TCA) using: (1) self-reported phenotypic predictors; (2) PGS predictors; and (3) combined models. Backward stepwise selection identified parsimonious predictor sets, with model fit assessed using AIC and likelihood ratio tests. Smoking status, sex, and core MDD symptoms were excluded due to missing data, extreme imbalance, or quasi-complete separation. All quantitative predictors were standardised (mean=0, SD=1).

#### GWAS on SSRI and SNRI Sustained Use and Self-Reported Efficacy

The primary GWAS compared participants with SSRI or SNRI acceptability (cases, n=5,554) versus non-acceptability (controls, n=2,270). Cases were defined as participants assigned to the SSRI or SNRI treatment acceptability groups. Controls included participants not assigned to these treatment groups who also had no history of sustained use (≥360 days) of any SSRI or SNRI medication in the 4.5-year window and did not self-report BIP. A secondary analysis focused specifically on SSRI acceptability (cases n=3,423, controls n=4,561).

As a complementary approach, we performed GWAS analyses using self-reported efficacy data for 10 commonly prescribed antidepressants. For the SSRI or SNRI efficacy phenotype, cases (n=8,223) self-reported positive response ("Moderately" to "Very well") to at least one SSRI or SNRI medication, while controls (n=904) did not self-report a positive response ("Not at all well") to any SSRI or SNRI but had available efficacy data for at least one of the 10 antidepressants studied. Participants with missing data for all medications were excluded. A secondary analysis examined SSRI-only efficacy (cases n=6,474, controls n=2,653).

All GWAS analyses were performed using PLINK 2.0 with adjustment for age, sex, and the first three genetic principal components. Only participants of genetically-inferred European ancestry were included, and individuals related at the second-degree or closer were removed (PLINK 2.0 --king-cutoff 0.0884). LD clumping identified independent loci using: significance threshold P < 5×10⁻⁸, LD threshold r²=0.1, and distance threshold of 250kb (but report all SNPs that surpassed suggestive significance, p < 5.0 x ${10}^{-6},$ in the supplementary material). SNP-based heritability on the observed scale were estimated using SBayesRC with GCTB software using ~7.4 million SNPs.

### Results

#### Supplementary Material 1: Australian Guideline Antidepressant Coverage

The following provides additional context on how our study medications align with Australian prescribing guidelines. Of the 7 “Choice” antidepressants recommended by the 2020 Australian prescribing guidelines^8^, four were included as study medications: escitalopram (SSRI), venlafaxine (SNRI), mirtazapine (TeCA) and amitriptyline (TCA). These medications correspond to four classes within the NbN2 neurobiology-based classification system (A11a1, A11b, AIII, and AIVc respectively)^9^, though we use the traditional classifications throughout our analysis. Two "Choice" medications (vortioxetine and agomelatine) were not common in the AGDS cohort, as they have been marketed only recently^45^ and the guidelines were published three years after the end of study follow-up^10^. Interestingly, bupropion, the last “Choice” medication has not yet been approved for depression treatment in Australia.

#### Supplementary Material 2: Phenotypic Factors Associated with Treatment Complexity

Half of the 40 tested self-reported traits were significantly associated with total AD dispense after Bonferroni correction, and an even larger subset was associated with AD medication or class diversity. (**Fig. 2, Supp. Table 3**). Only a few traits (T2D, back pain, and feelings of guilt) were associated with greater dispense but not increased AD class or medication diversity, suggesting these characteristics are associated with prolonged AD use but not with DTD inferred from AD diversity. In contrast, traits such as regular smoking, increased alcohol consumption, lower education level, stomach ulcers, chronic fatigue syndrome (CFS), endometriosis, uterus fibroids, schizophrenia, substance use disorder (SUD), and specific MDD symptoms (e.g., appetite/weight changes and death thoughts) were associated with greater treatment diversity but not overall dispense. These findings align with prior evidence that higher education is associated with lower rates of antidepressant switching^11^ and a reduced risk of TRD^12^, and with evidence that smoking can decrease antidepressant efficacy through pharmacokinetic interactions^13^. Finally, traits significantly associated to both greater dispense and diversity, including recurrent MDD, suicidal ideation, self-harm, poor subjective physical health, higher BMI, migraines/headaches, chronic pain, personality disorder (PersD), and core MDD symptoms such as sleep disturbances and movement changes, imply a more severe depression or comorbid clinical profiles requiring both long-term care and frequent pharmacological adjustments, hallmark features of DTD^14^.

#### Supplementary Material 3­­: Pharmaceutical Dispensing Patterns by Treatment Group

We first investigated whether dispensing patterns varied meaningfully across medication subgroups (**Fig. 4, Supp. Table 9-10, Supp. Fig 4**). A clear SNRI class effect occurred, with SNRI sustained users having significantly higher cumulative medication dispensing compared to SSRI sustained users (β = 0.23, SE = 0.018, p = 9.90 x${10}^{-35}$) alongside greater antidepressant class diversity (β = 0.21, SE = 0.025, p = 7.50 x ${10}^{-17}$), indicative of sequential treatment attempts and a longer therapeutic trajectory. SNRI sustained use was also associated with extended prescription episode durations (β = 0.14, SE = 0.024, p = 6.20 ×${10}^{-9}$) and fewer prescription episodes (β = –0.10, SE = 0.021, p = 1.80 ×${10}^{-6}$), suggesting consistent use during efforts to achieve and maintain remission. The estimate of class diversity relative to the SSRI group was highest for the TeCA (i.e., mirtazapine) sustained use group (β = 0.41, SE = 0.07, p = 2.70 × ${10}^{-9}$), consistent with sequential pharmacological trials, combination strategies, and potential TRD. In contrast, while the TCA (i.e., amitriptyline) group also showed elevated rates of medication class diversity (β=0.33, SE=0.076, p=1.00 x${10}^{-5}$), this group was the only one characterized by markedly shorter individual prescription episodes (β = –0.29, SE = 0.073, p = 7.60 x ${10}^{-5}$), while having a significantly greater number of prescription episodes (β = 0.25, SE = 0.063, p = 9.30 x ${10}^{-5}$), consistent with frequent, shorter episodic prescribing patterns.

#### Supplementary Material 4: Participant Phenotypes Differentially Associate with Choice of Antidepressant Medication

Building on the dispensing patterns, we next explored clinical characteristics and comorbidities associated with sustained antidepressant use with class effect size estimates expressed relative to SSRIs (**Supp. Tables 9-12, Supp. Fig. 4-8**).

##### SNRIs

SNRI users had significantly higher BMI (β = 0.67, SE = 0.19, p = 4.0 × ${10}^{-4}$), a pattern consistent across individual SNRIs: duloxetine (β = 1.35, SE = 0.38, p = 3.5 × ${10}^{-4}$), desvenlafaxine (β = 0.92, SE = 0.33, p = 0.005) and venlafaxine (β = 0.75, SE = 0.32, p = 0.019), all relative to sertraline. While long-term antidepressant use is linked to weight gain^15-17^, large-scale U.S. EHR data have reported only small differences between SSRIs and SNRIs^18^. However, that study also found slightly greater weight gain with duloxetine versus sertraline^18^, consistent with our findings. Among SNRIs, duloxetine was associated with the lowest subjective physical health scores (β = -0.18, SE = 0.057, p = 1.4 × ${10}^{-3}$), mirroring its association with higher BMI. SNRI users also reported more MDD symptoms during their worst episodes, including increased rates of depressed mood (OR = 2.4, CI = 1.6–3.6, p = 2.1 × ${10}^{-5}$) and anhedonia (OR = 1.8, CI = 1.4–2.3, p = 1.5 ×${10}^{-6}$).

Consistent with this severity profile, SNRI users had a higher lifetime psychiatric comorbidity burden, particularly for PersD (OR = 1.7, CI = 1.3–2.2, p = 3.7 ×${10}^{-5}$) and SUD (OR = 1.5, CI = 1.1–2.0, p = 0.015), reflecting the well-established co-occurrence of MD, PersD and SUD^19^. However, SNRI users were less likely to self-report comorbid OCD (OR = 0.68, CI = 0.52–0.88), potentially reflecting prescriber patterns favouring SSRIs for OCD^20^. Duloxetine showed strong associations with somatic comorbidities, including chronic pain (OR = 2.2, CI = 1.7–3.0, p = 1.2 × ${10}^{-7}$), back pain (OR = 1.4, CI = 1.1–1.7, p = 0.0067), and chronic fatigue syndrome (OR = 1.8, CI = 1.2–2.7, p = 0.0045), consistent with its use as an analgesic^21^ and the broader clinical picture of poorer subjective physical health in this group. Paradoxically, despite having the highest self-report BMI, the duloxetine group showed elevated rates of self-report lifetime anorexia nervosa (AN, OR = 2.3, CI = 1.3-3.9, p = 3.7 x ${10}^{-3}$) relative to sertraline, highlighting the complex nature of eating disorder presentations.

##### TCAs (Amitriptyline)

TCA users were significantly older (β = 8.7, SE = 1.2, p = 2.6 × ${10}^{-12}$) with lower education attainment (β = –0.32, SE = 0.083, p = 1.2 ×${10}^{-4}$) and the lowest physical health scores overall (β = –0.36, SE = 0.09, p = 8.5 ×${10}^{-5}$), despite similar BMI to SSRI users. Notably, TCA use was uniquely associated with reduced depressed mood relative to SSRIs (OR = 0.36, CI = 0.2-0.62, p = 2.9 x ${10}^{-4}$), suggesting utility in MDD with comorbid somatic conditions. This pattern was reinforced by their comorbidity profile, showing significantly more somatic conditions including T2D (OR = 2.1, CI = 1.2–3.6, p = 0.0074), chronic pain (OR = 4.2, CI = 2.9–5.9, p = 5.3 × ${10}^{-15}$), migraines (OR = 1.8, CI = 1.2–2.9, p = 0.0054), and endometriosis (OR = 2.1, CI = 1.2–3.4, p = 0.0054). This somatic profile is consistent with their use in migraine prevention^22^ and neuropathic pain management^23^. Importantly, TCA users did not show elevated psychiatric comorbidities, distinguishing them from other treatment groups.

##### TeCAs (Mirtazapine)

Similar to the TCA group, relative to the SSRI sustained use group, TeCA users were older (β = 5.54, SE = 1.2, p = 1.5 × ${10}^{-6}$) with lower education attainment (β = –0.29, SE = 0.077, p = 1.2 ×${10}^{-4}$). Male overrepresentation was notable in the TeCA group (OR = 3.1, CI = 2.3–4.2, p = 6.6 ×${10}^{-13}$). They had higher rates of suicidal ideation (OR = 1.8, CI = 1.3–2.6, p = 8.7 ×${10}^{-4}$) and death thoughts (OR = 2.0, CI = 1.3–3.2, p = 0.0020), likely reflecting mirtazapine’s preferential use in more severe or TRD^24^, and its faster onset of action compared to SSRIs^12,25^. At the drug-level, mirtazapine users showed increased appetite/weight change (OR = 1.9, CI = 1.2–3.1, p = 0.0019), consistent with its known appetite-stimulating effects^12,25^ via 5-HT(3) blockade^26^. Although mirtazapine has shown to benefit insomnia via circadian resynchronisation^27^, and in reducing suicidality in insomnia patients^28^, we found no evidence of increased sleep disturbances during peak depressive episodes. Unlike TCA users, TeCA users showed significantly higher rates of PersD (OR = 1.8, CI = 1.3–2.6, p = 8.7 × 10⁻⁴), but did not exhibit the somatic comorbidity pattern seen with TCAs, aligning with evidence that TCAs are less effective in individuals with comorbid PersD^29^.

##### Self-report Bipolar Disorder Groups (BIP+/-L)

Both groups of participants living with bipolar disorder experienced more depressive episodes than SSRIs users (BIP-L: β = 0.6, SE = 0.038, p = 5.9 × ${10}^{-54}$, BIP+L: β = 0.55, SE = 0.07, p = 5.9× ${10}^{-15}$), and had higher self-reported BMI (BIP-L: β=1.39, SE=0.27, p=2.7 x ${10}^{-7}$; BIP+L: β=1.59, SE=0.49, p=1.2 x ${10}^{-3}$). Unlike the BIP+L group, the BIP-L group reported significantly more mood-related symptoms after Bonferroni correction – low-interest (OR = 5.4, CI = 2.0-9.7, p = 1.7 x ${10}^{-8}$), depressed mood (OR=4.6, CI=2-11, p=2.7 x ${10}^{-4}$) and guilty feelings (OR=2.1, CI=1.4-3.1, p=2.0 x ${10}^{-4}$). Whereas the BIP+L group, compared to the BIP-L group, had a nominally higher rate of the neurovegetative symptom, fatigue (OR=8.8, CI=1.2-65, p=3.2 x${10}^{-2}$).

The BIP-L group showed the broadest psychiatric comorbidity profile, with highest odds of PersD (OR = 5.4, CI = 4.2–7, p = 1.7 × ${10}^{-37}$), ADHD (OR = 3.2, CI = 2.3-4.4, p = 2.2 x ${10}^{-13}$), seasonal affective disorder (SAD; OR = 3, CI = 2.1-4.2, p = 3.4 x ${10}^{-10}$), OCD (OR = 1.9, CI = 1.4-2.4, p = 3.5 x ${10}^{-6}$), and uniquely, premenstrual dysphoric disorder (PMDD; OR = 2.2, CI = 1.5-3.2, p = 7.7 x ${10}^{-5})$. Whilst also showing association with lower education attainment (β = –0.18, SE = 0.038, p = 2.6 × ${10}^{-6}$) and earlier depression onset (β = –0.28, SE = 0.033, p = 1.9 × ${10}^{-17}$). In contrast, the BIP+L group showed a severe depression profile, with the highest rate of suicidal ideation (OR = 4.2, CI = 2.8–6.4, p = 1.0 × ${10}^{-11}$) consistent with lithium often being reserved for severe depression in BIP and has been associated with reduced suicide risk following recent self-harm^30^. The group also had a more focused comorbidity profile enriched for SUD (OR = 5.6, CI = 3.5-8.8, p = 1.9 × ${10}^{-13}$), schizophrenia (OR = 5.0, CI = 2.2-11, 9.8 x ${10}^{-5}$), PersD (OR = 3.7, CI = 2.3–5.8, p = 2.8 × ${10}^{-8}$) and ADHD (OR = 2.7, CI = 1.6-4.7, p = 2.4 x ${10}^{-4}$). These findings suggest that the BIP−L group captures a more heterogenous subset characterised by extensive psychiatric comorbidity and earlier depression onset. Moreover, the narrower comorbidity profile in the BIP+L group is consistent with literature showing that lithium response is associated with fewer comorbidities than with non-response^31^.
