## Supplementary Figures for "Decoding Treatment Choice: Genetic and Phenotypic Analyses of Long-term Antidepressant Acceptability"

**Supplementary Figure 1.**

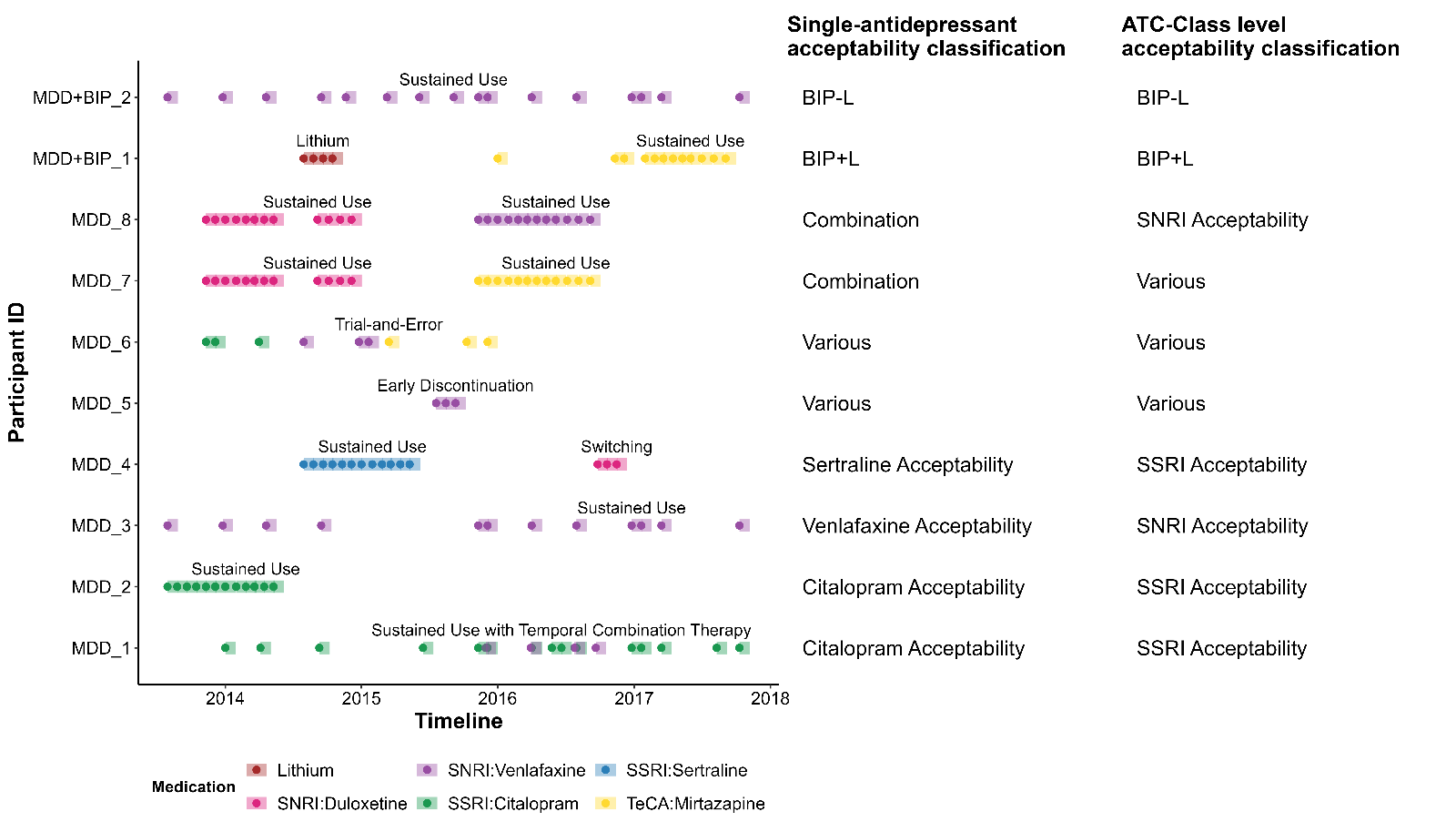
**Sustained use threshold as a proxy for antidepressant acceptability and treatment group classifications.** Column 1: Example patterns of antidepressant use over a 4.5-year period. Column 2: Corresponding single-antidepressant acceptability classification. Column 3: Corresponding class-level acceptability allocation (i.e., SSRI, SNRI, TeCA, TCA, BIP-L, BIP+L, Various). To approximate treatment duration using PBS dispensing data, we first applied a standard 30-day duration per dispense based on typical Australian prescribing practices (Step 1). To avoid overestimating treatment duration due to overlapping prescriptions, the per-prescription duration was calculated as the minimum of either (a) the time until the next dispense of the same medication, or (b) 30 days (Step 2). Cumulative duration was then calculated by summing individual prescription durations, allowing for natural gaps in treatment (Step 3). Treatment acceptability was defined as sustained cumulative use (≥360 days) of a single antidepressant. This proxy does not account for strict adherence or continuous use. Cases involving intermittent patterns—such as medication cessation and re-initiation or dispensing in sporadic <8-week blocks—may still be classified as "acceptable" if the total cumulative duration exceeds the 360-day threshold over the 4.5-year window (e.g., MDD_1, MDD_3).

**Supplementary Figure 2**

**Antidepressant use and outcomes in the Australian Genetics of Depression Study (AGDS).** Data are shown for 13,763 AGDS participants who had ≥1 recorded dispense for one of the 10 most commonly prescribed antidepressants in the cohort, based on linked Pharmaceutical Benefits Scheme (PBS) records from July 2013 to December 2018. These antidepressants are grouped by class (SSRI, SNRI, TeCA, TCA) and summarized across five panels: (A) Number of participants with at least one recorded dispense for each class. (B) Distribution of cumulative prescription duration per antidepressant within each class. The dashed line indicates the threshold used to define antidepressant acceptability (≥ 360 days). (C) Cumulative prescription duration relative to SSRIs (used as the reference class), estimated using linear regression adjusted for age and sex (N = 13,693). The marginal mean for SSRI is shown in black. Classes with statistically significant differences are shown in blue; non-significant differences are shown in grey. (D) Proportion of participants reporting a positive treatment response to an AD, summarised by class stratified by increasing thresholds of cumulative prescription duration (range: ≥1 to ≥1500 days). Responses of “Moderately well” or “Very well” were classified as responders; “Not at all” as non-responders. The dashed line indicates the acceptability threshold. (E) Proportion of participants reporting discontinuation due to any side effects, summarised by class and stratified by cumulative prescription duration (range: ≥1 to ≥1500 days), with the same dashed line indicating the acceptability threshold.

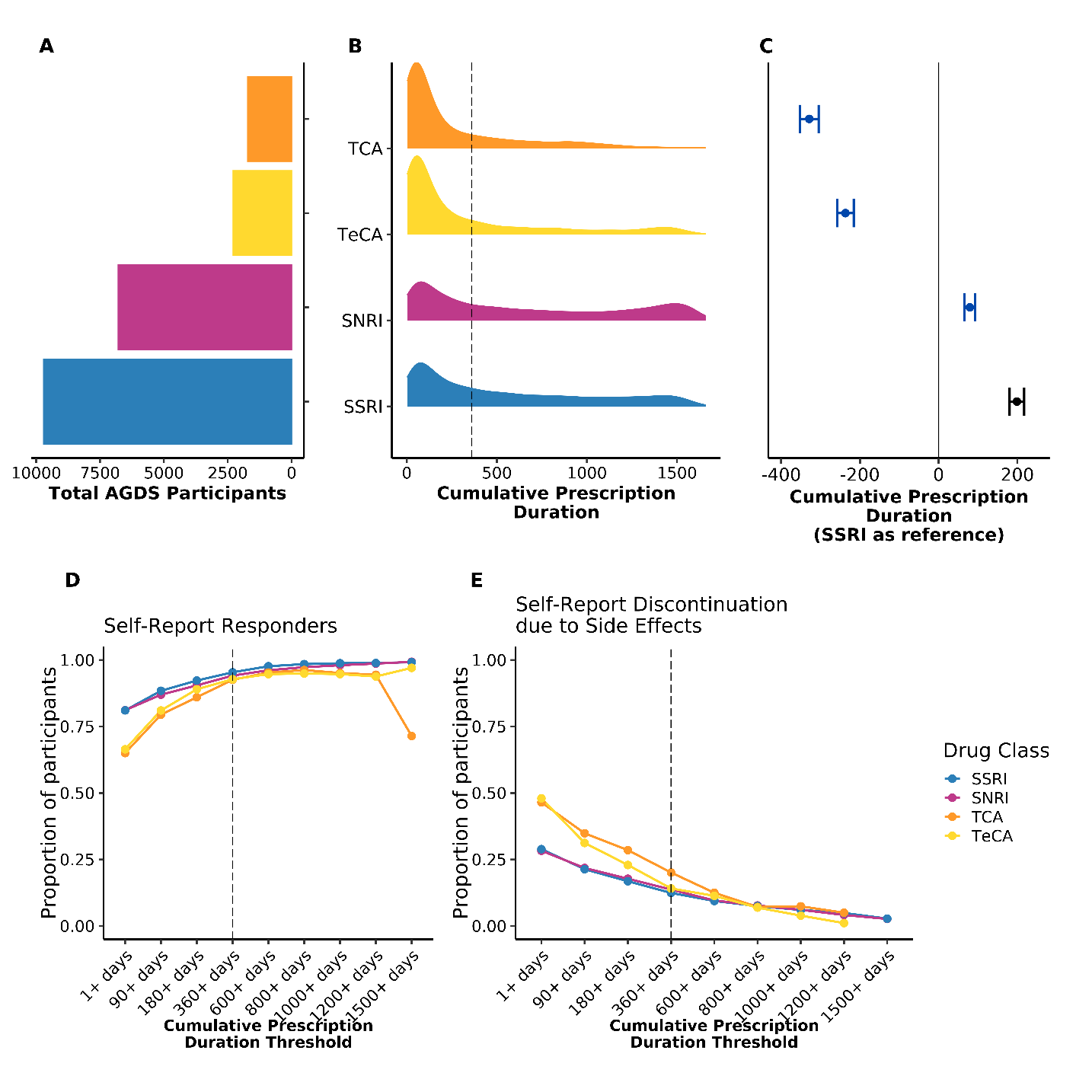

**Supplementary Figure 3.**

**Stable rates of recurrent depressive and prescription episodes across cumulative prescription duration threshold**. This figure summarizes trends in recurrent episodes across different levels of cumulative prescription exposure: (A) Rates of repeat prescription episodes, defined as new antidepressant dispensing events separated by gaps of more than 90 days (i.e., independent treatment episodes). (B) of self-reported lifetime depression episodes. (C) Median total prescription duration (TPD), with interquartile range (IQR) estimates, plotted across total prescription episodes (TPE) for individuals with less than 800 days of cumulative TPD. Median TPD generally increases with a higher number of TPEs. The apparent decline in repeat prescription episodes beyond approximately 600–800 days of cumulative TPD (panel A) is likely an artifact of the cross-sectional data window rather than a therapeutic effect. Longer treatment durations reduce the opportunity for observing additional independent episodes within the fixed observation period.

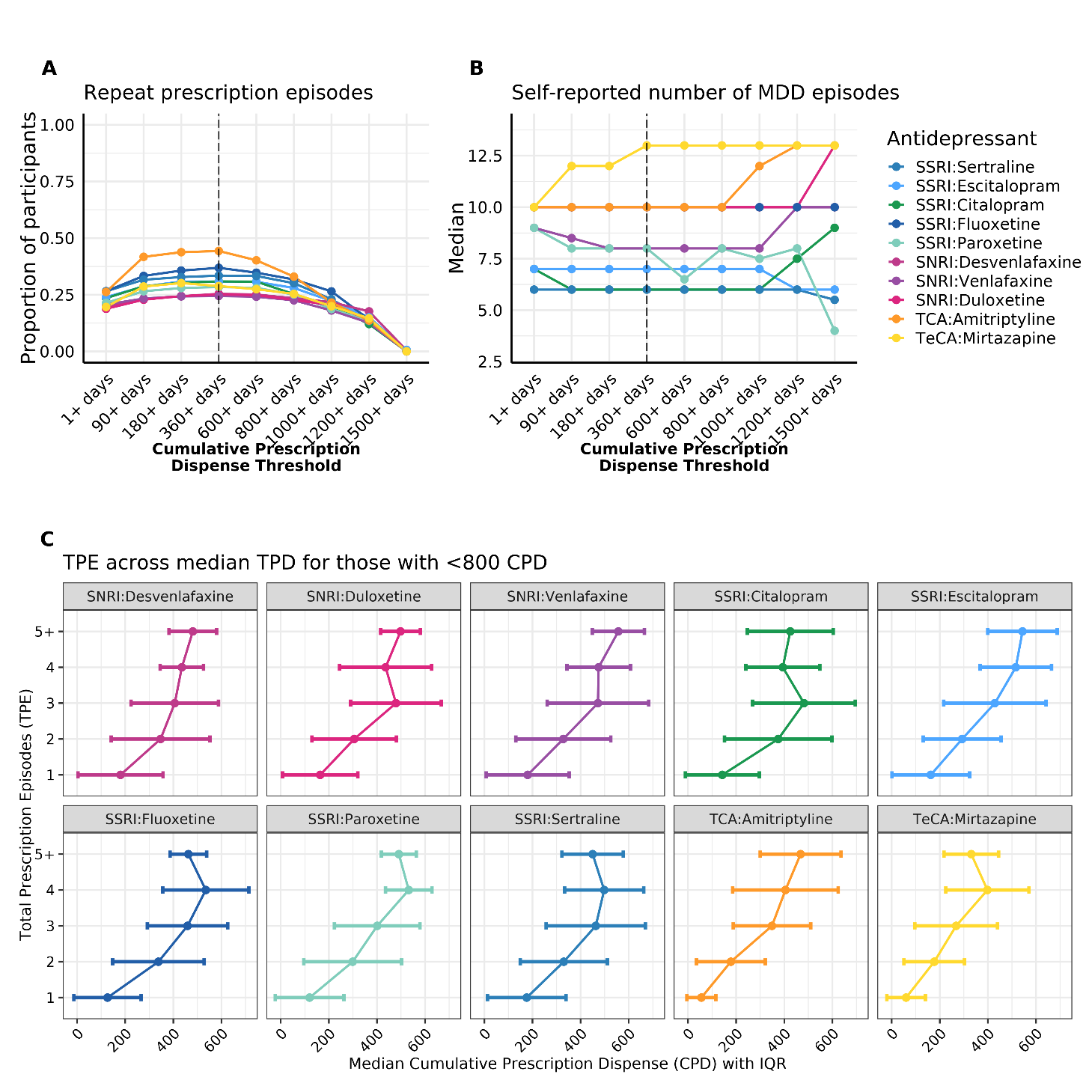

**Supplementary Figure 4.**

**Associations between 40 self-reported traits and 3 antidepressant (AD) use metrics —cumulative AD dispense, medication diversity, and class diversity—among 13,763 AGDS participants.** (1) Left panel: Cumulative all antidepressant prescription duration (days); (2) Middle panel: AD diversity (range: 1–10); (3) Right panel: Class diversity (range: 1–4). Associations were adjusted for age and sex, except for when these were the variables of interest. Statistical significance was declared at p < 0.05 after false discovery rate (FDR) correction (*) and Bonferroni correction (**), applied separately within each DTD metric. The y-axis reports the number of AGDS participants with complete data for age, sex and the binary trait, as well as the % of cases. For interpretation: higher subjective physical health scores reflect better health; higher education levels reflect greater educational attainment; female is the reference group in sex. See Supp. Table 3 for complete results.

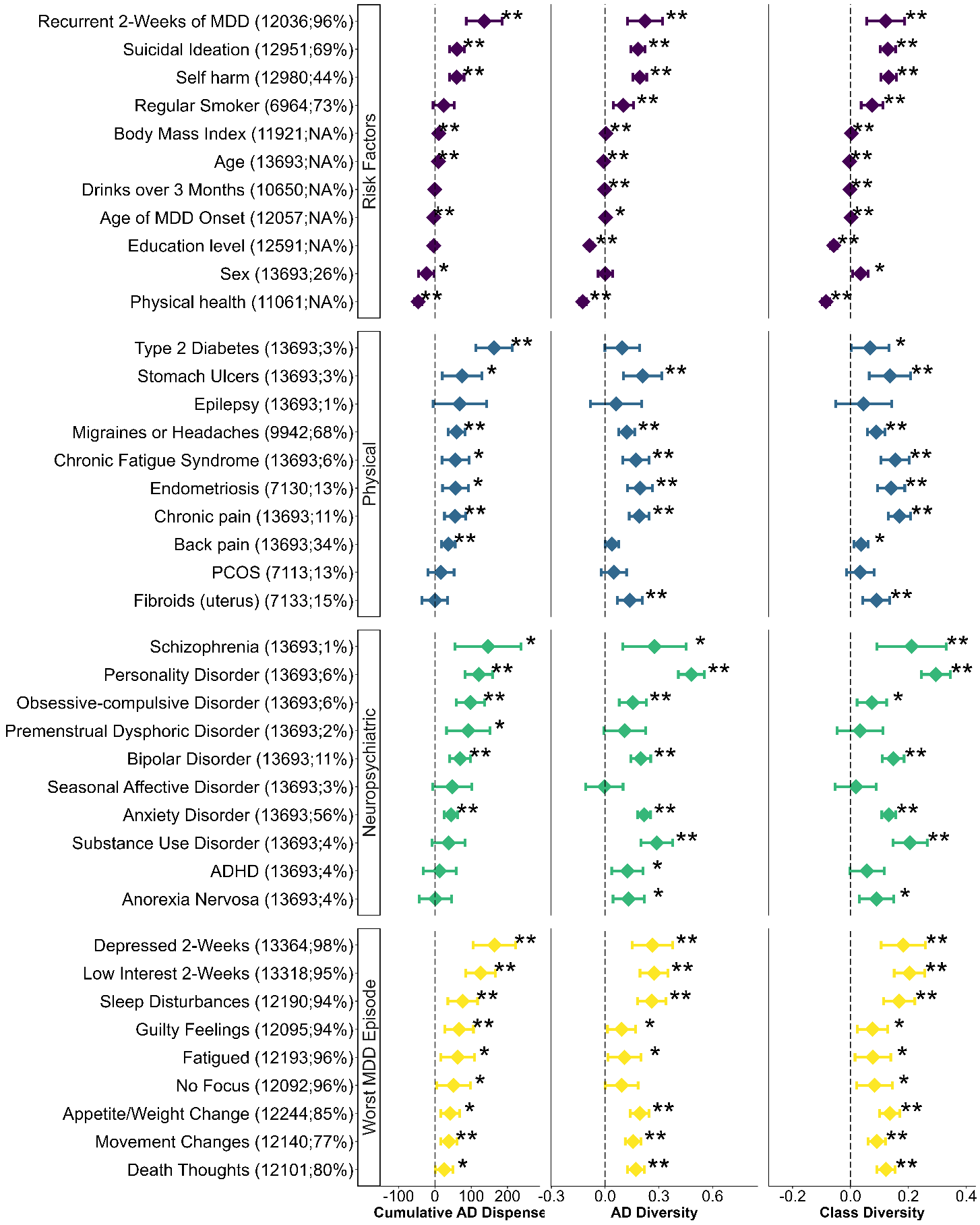

**Supplementary Figures 5 and 6. Associations between self-reported quantitative variables and antidepressant acceptability under two sustained use thresholds.** Analyses were conducted separately under two cumulative dispensing thresholds: (1) ≥360 days and (2) ≥600 days. Supplementary Figure 5 presents associations with pharmaceutical metrics. Supplementary Figure 6 presents associations with clinical features. All variables, except age and BMI, were standardized (mean = 0, SD = 1) across the full AGDS cohort with at least one recorded antidepressant dispense (N = 13,763). Linear models were adjusted for age (in years) and sex (female as reference), except when age was the dependent variable. Statistical significance was defined as *p* < 0.05 following multiple testing correction. Asterisks indicate significance after false discovery rate (FDR) correction (*) and Bonferroni correction (**), applied separately for each threshold–drug group combination.

**Supplementary Figure 5**

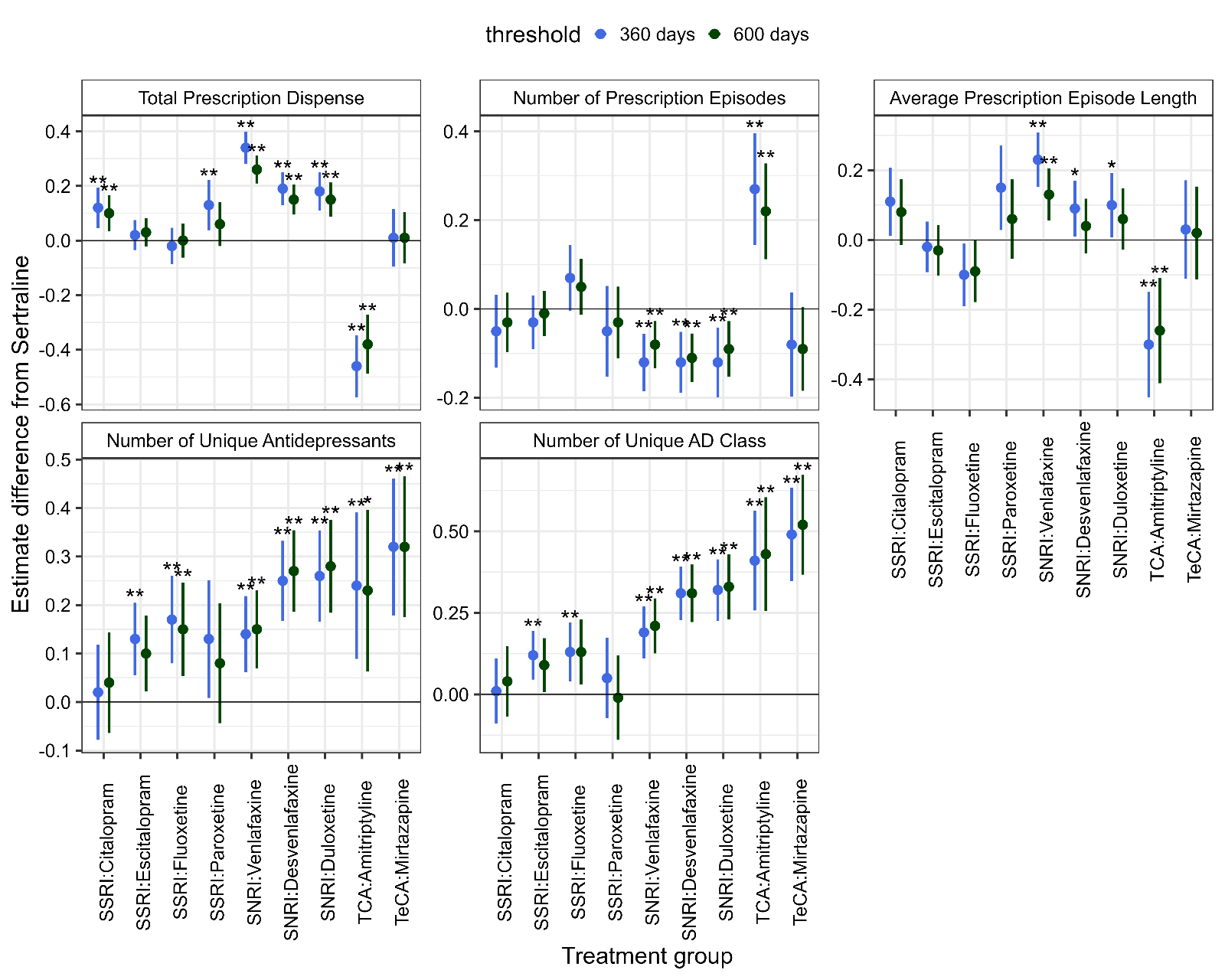

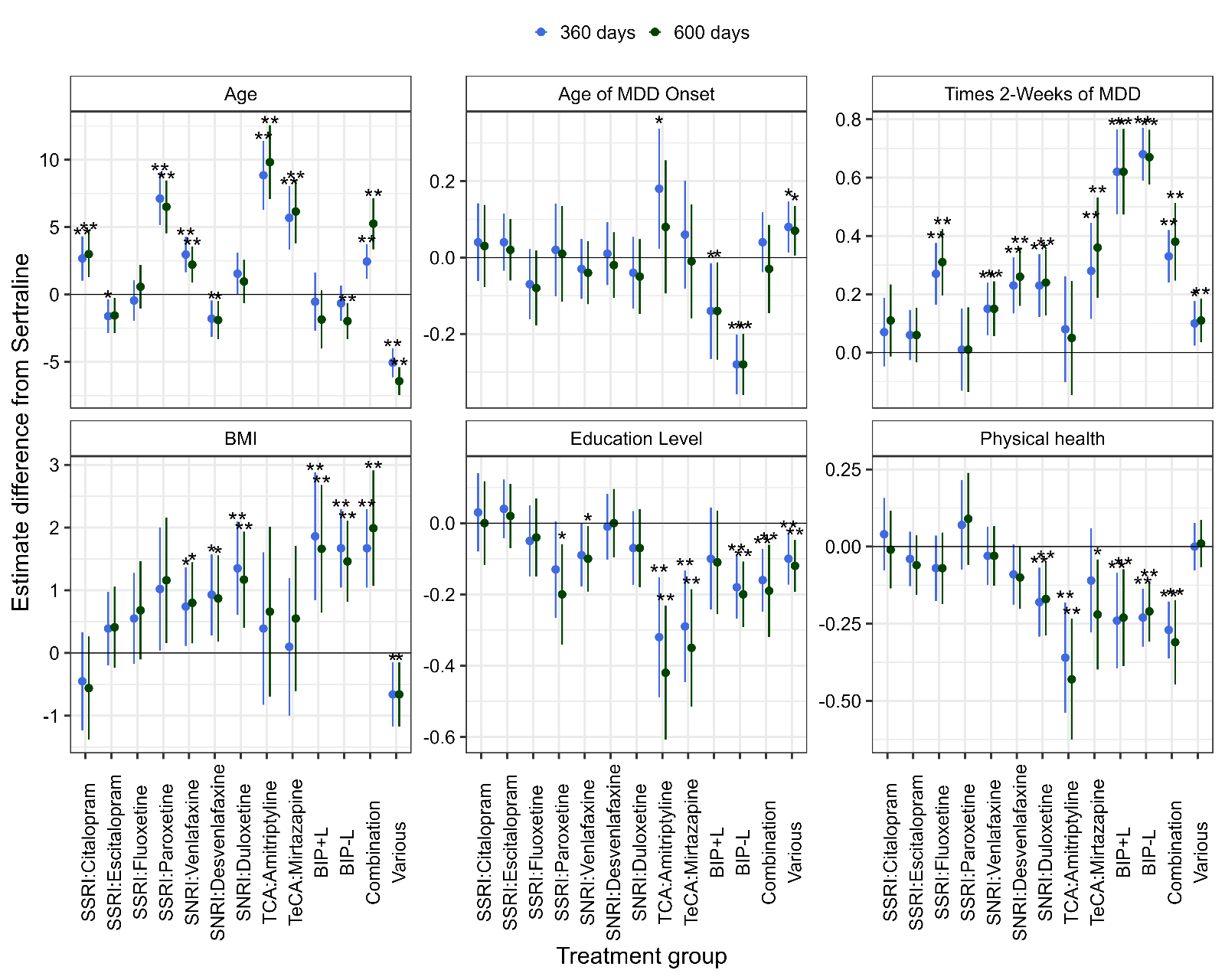
**Supplementary Figure 6**

**Supplementary Figures 7, 8 and 9. Associations between self-reported binary variables and antidepressant acceptability under two sustained use thresholds.** Analyses were conducted separately under two cumulative dispensing thresholds: (1) ≥360 days and (2) ≥600 days. Supplementary Figure 7 presents associations with risk factors and treatment response phenotypes. Supplementary Figure 8 presents associations with physical and psychiatric traits. Supplementary Figure 9 presents associations with symptoms reported during the worst MDD episode. All variables were standardized (mean = 0, SD = 1) across the full AGDS cohort with ≥1 antidepressant dispense (N = 13,763). Models were adjusted for age (in years) and sex (female as reference), except when sex was the outcome. Statistical significance was defined as p < 0.05 after false discovery rate (FDR) correction (*) and Bonferroni correction (**), applied separately within each threshold-drug group combination.

**Supplementary Figure 7**

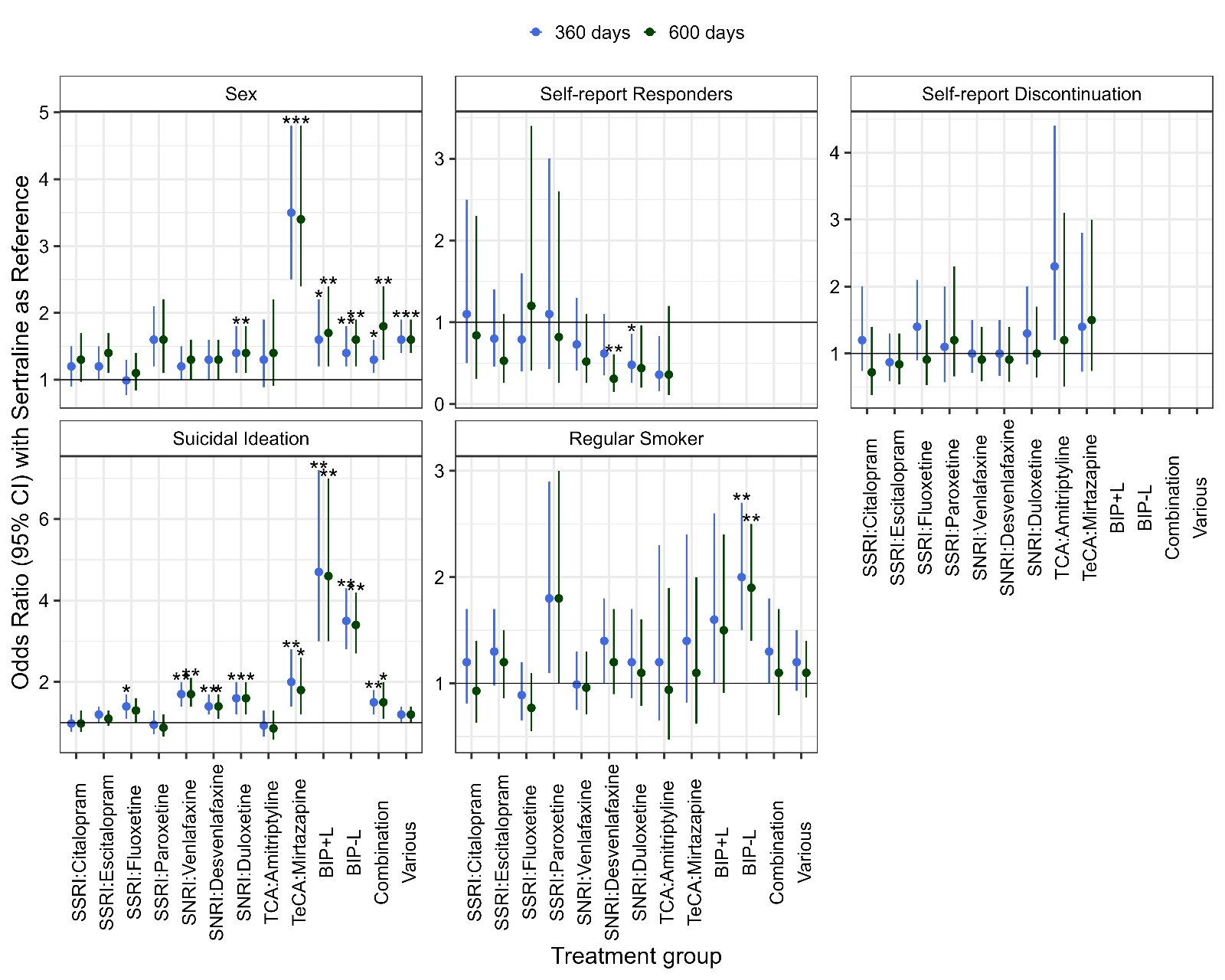

**Supplementary Figure 8**

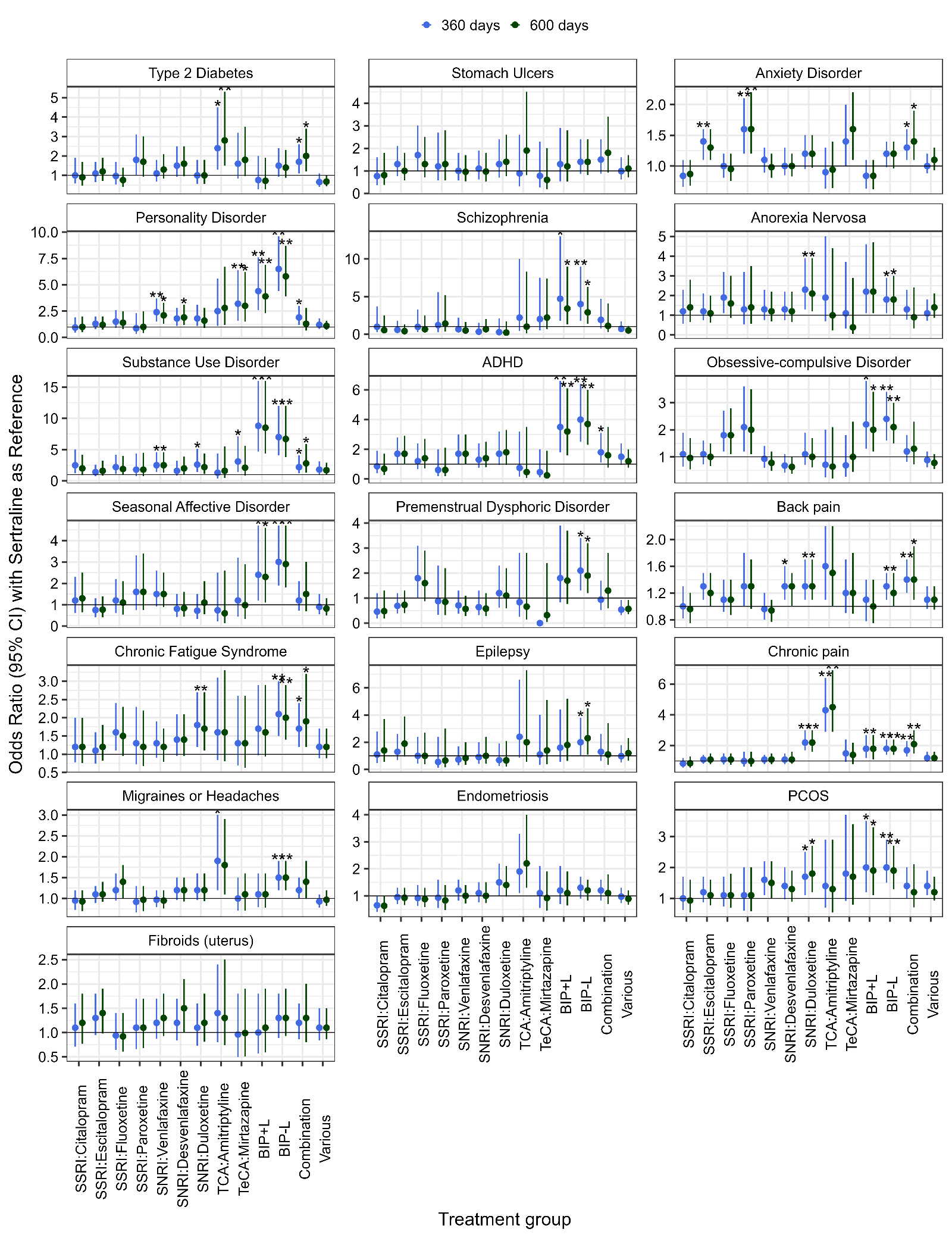

**Supplementary Figure 9**

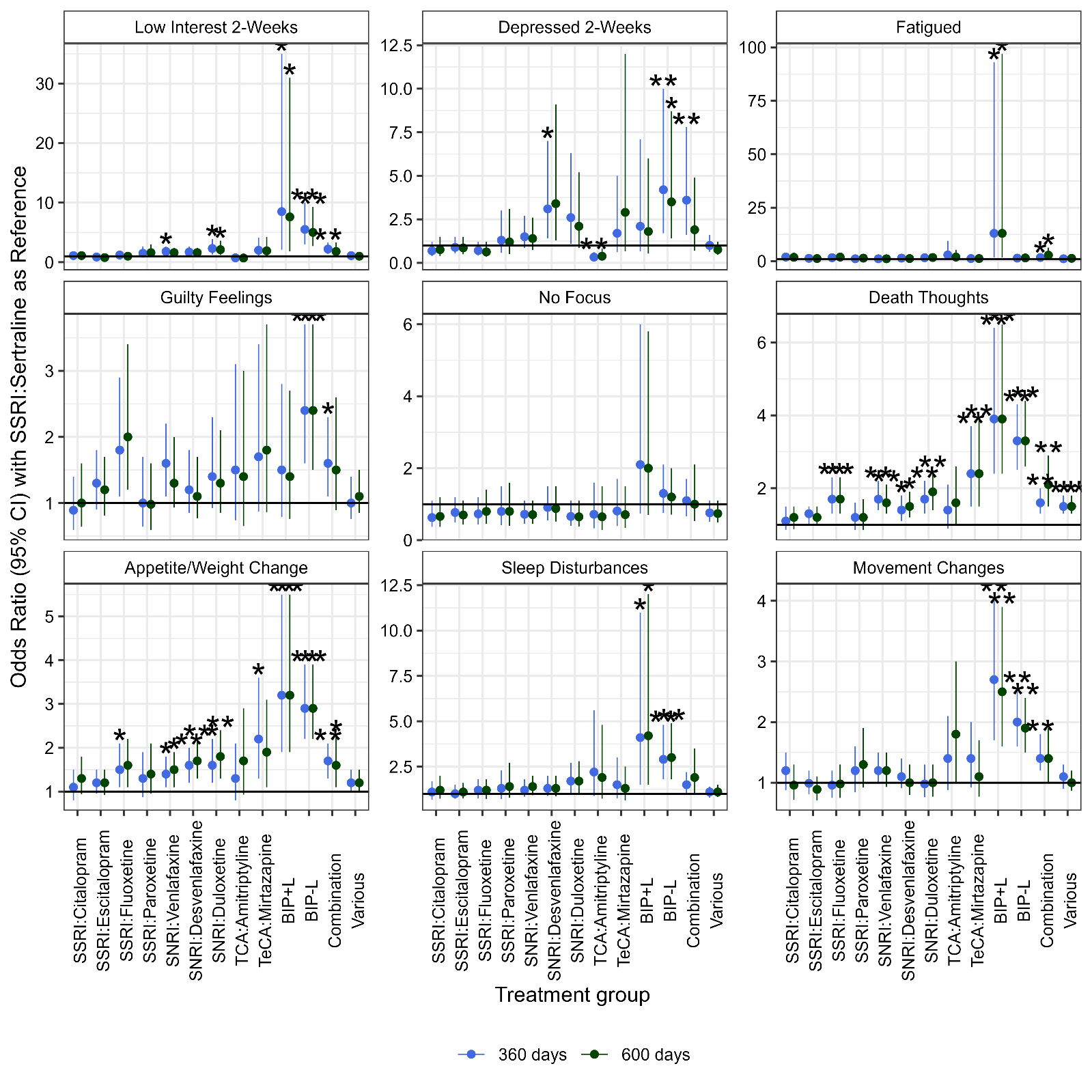

**Supplementary Figure 10**

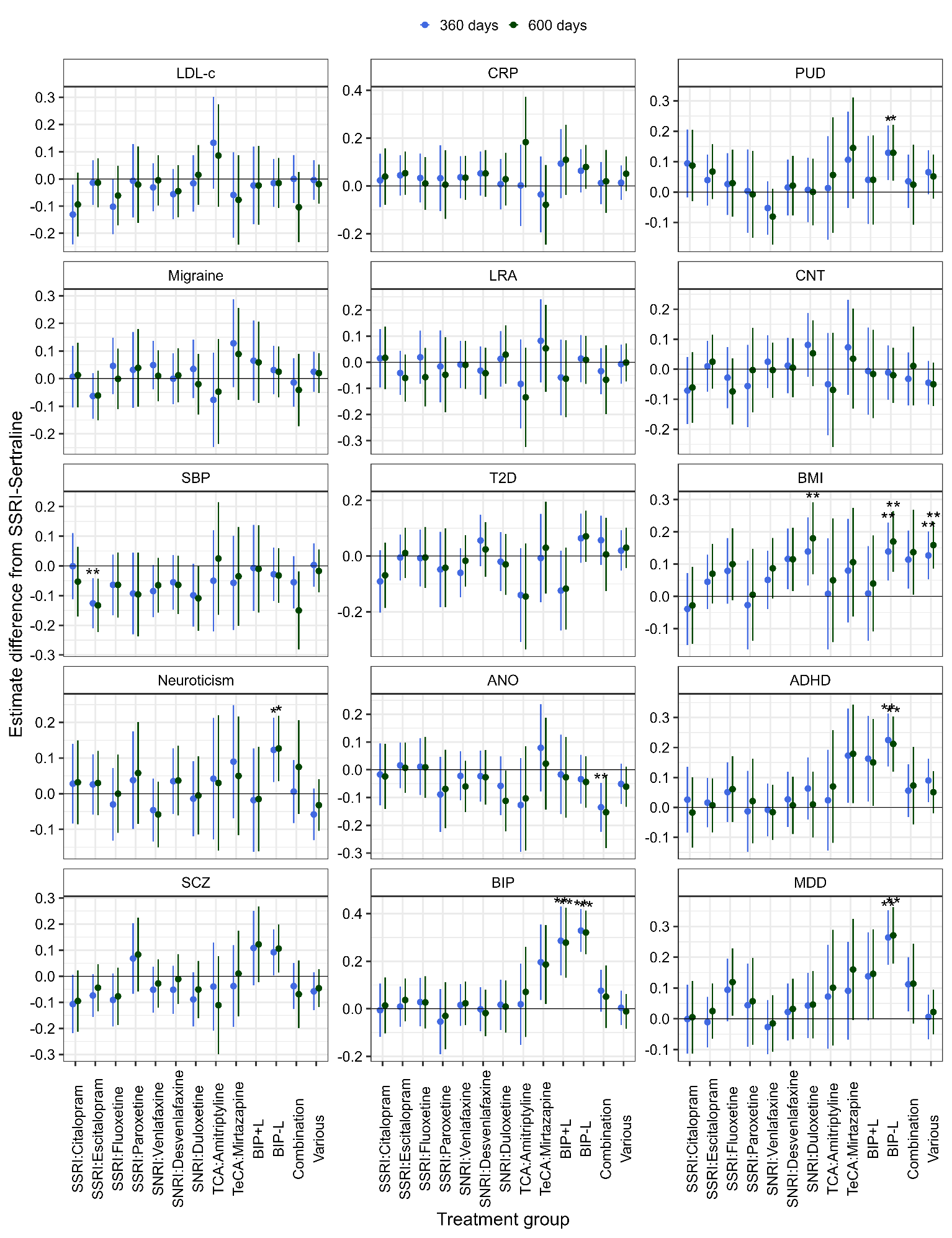
**Associations between polygenic risk scores and antidepressant acceptability under two sustained use thresholds.** Associations between 15 polygenic risk scores (PGS) and drug-level antidepressant acceptability groups were examined separately under two cumulative dispensing thresholds: (1) ≥360 days and (2) ≥600 days. All PGS were standardized across 14,603 AGDS participants of genetically inferred European ancestry. Statistical significance was defined as p < 0.05 following multiple testing correction. Asterisks indicate significance after false discovery rate (FDR) correction (*) and Bonferroni correction (**), applied separately within each threshold group.

**Supplementary Figure 11**

**Association between BMI and BMI polygenic score (PGS) with antidepressant acceptability under two sustained use thresholds.** Body mass index (BMI), calculated from self-reported height and weight, was examined in relation to drug-level antidepressant acceptability groups under two cumulative dispensing thresholds: Top panel: ≥360 days; Bottom panel: ≥600 days. Analyses were conducted separately for each threshold group using linear regression models adjusted for age (in years), sex (female as reference), and BMI polygenic score (PGS). The PGS was standardized across the full AGDS cohort of genetically inferred European ancestry (N = 14,603). Statistical significance was defined as p < 0.05 after multiple testing correction, applied separately for each threshold group. Asterisks indicate significance after false discovery rate (FDR) correction (*) and Bonferroni correction (**).

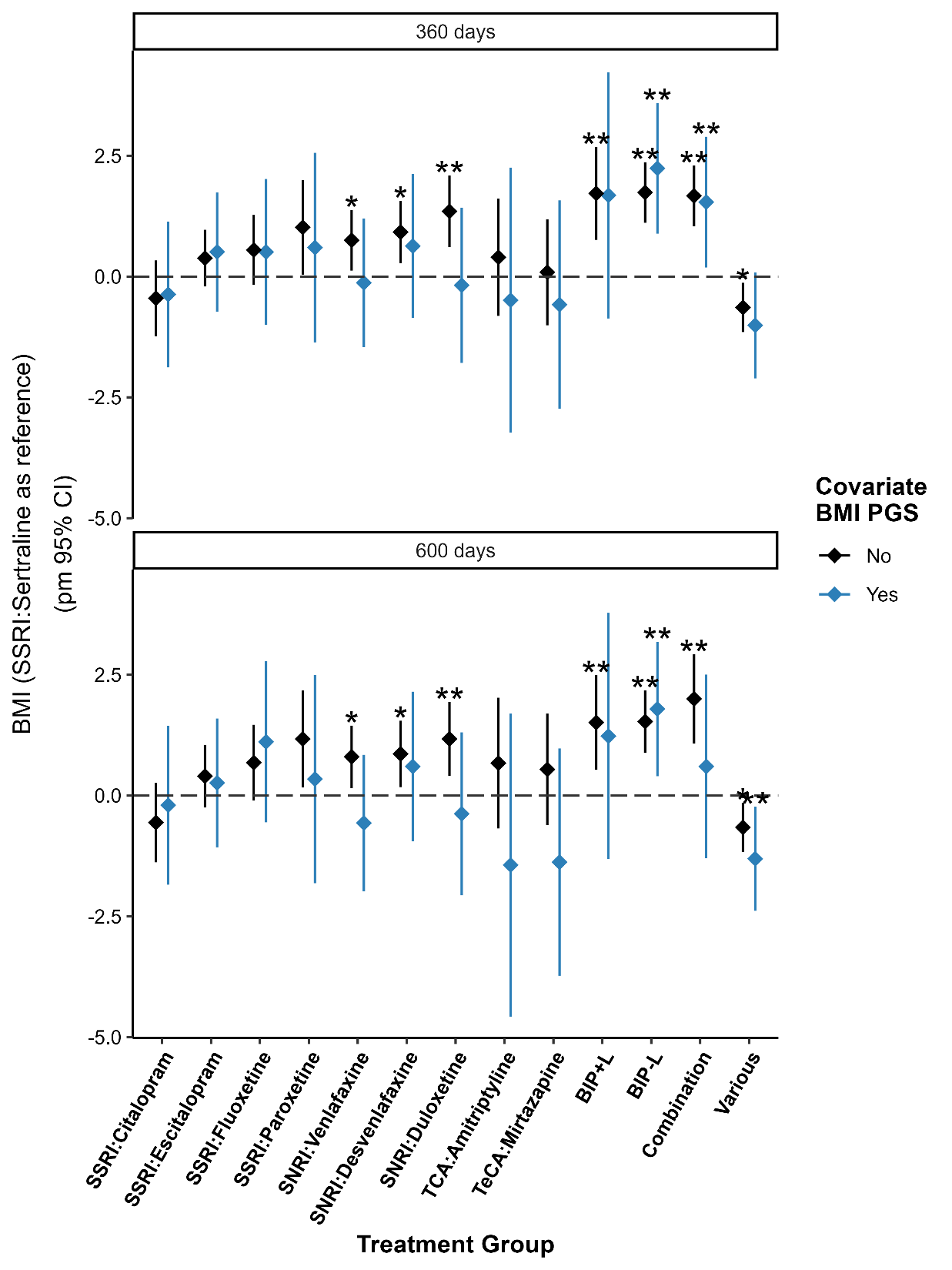

**Supplementary Figure 12**

**Multinomial logistic regression models predicting antidepressant class assignment**. Results are shown from three separate multinomial logistic regression models predicting antidepressant class (SSRI, SNRI, TeCA, TCA) based on: (A) self-reported phenotypic predictors; (B) polygenic score (PGS) predictors; and (C) a combined model including both phenotypic and PGS predictors. All quantitative predictors were standardized (mean = 0, SD = 1). For continuous predictors, odds ratios (ORs) represent the change in odds for a one standard deviation increase in the predictor. Backward stepwise selection was applied to each model to remove non-significant predictors and identify the most parsimonious final set of variables. The variable PDMD was excluded from the figure for visualization purposes.

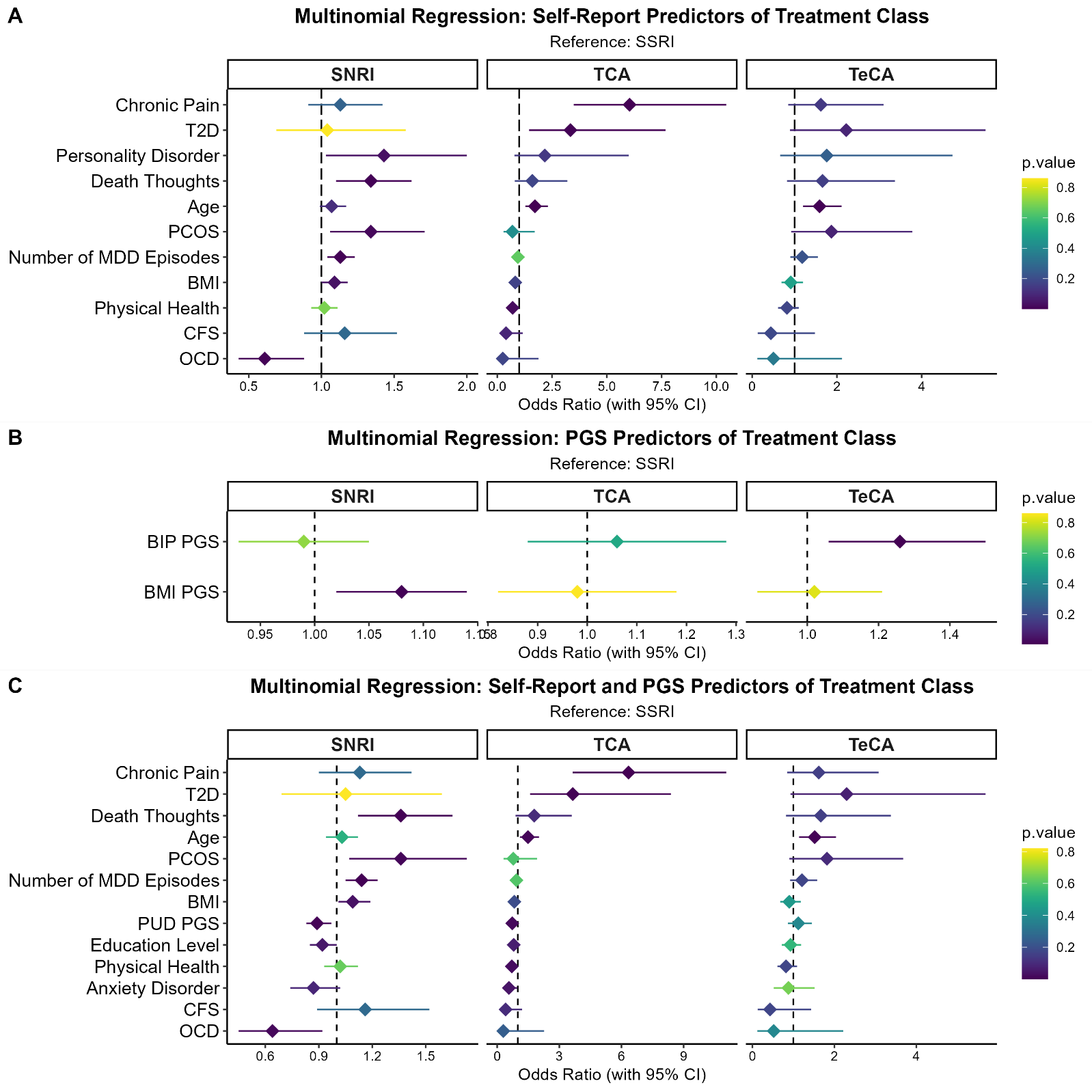

**Supplementary Figure 13 and 14. Genome-wide association study (GWAS) of SSRI and SNRI acceptability.** A logistic regression GWAS was conducted comparing participants with SSRI or SNRI acceptability (cases, n = 5,774) to those with non-acceptability (controls, n = 2,348). Controls were drawn from participants not classified as SSRI or SNRI acceptable, excluding those with sustained SSRI use (≥360 days) or self-reported bipolar disorder. Analyses were performed using PLINK 2.0, adjusting for age, sex, and the first three genetic principal components (PCs). Only participants of genetically inferred European ancestry were included, and individuals related at the second-degree or closer were removed (PLINK 2.0 --king-cutoff 0.0884). Supplementary Figure 13: Quantile-quantile (QQ) plot and the genomic inflation factor (λ = 0.997) indicating minimal inflation. Supplementary Figure 14: Manhattan plot of all common SNPs. SNPs surpassing suggestive significance (p < 5.0 × 10⁻⁶) are highlighted in green. SNP-based heritability was estimated using SBayesRC in GCTB software based on approximately 7.4 million SNPs. The SNP-based heritability was estimated at *h²* = 0.15 ± 0.160 (mean ± SD), based on an observed phenotypic variance of 0.21 for the binary SSRI/SNRI acceptability trait. The wide uncertainty reflects known limitations in heritability estimation for binary traits in moderately sized samples^1^

**Supplementary Figure 13**

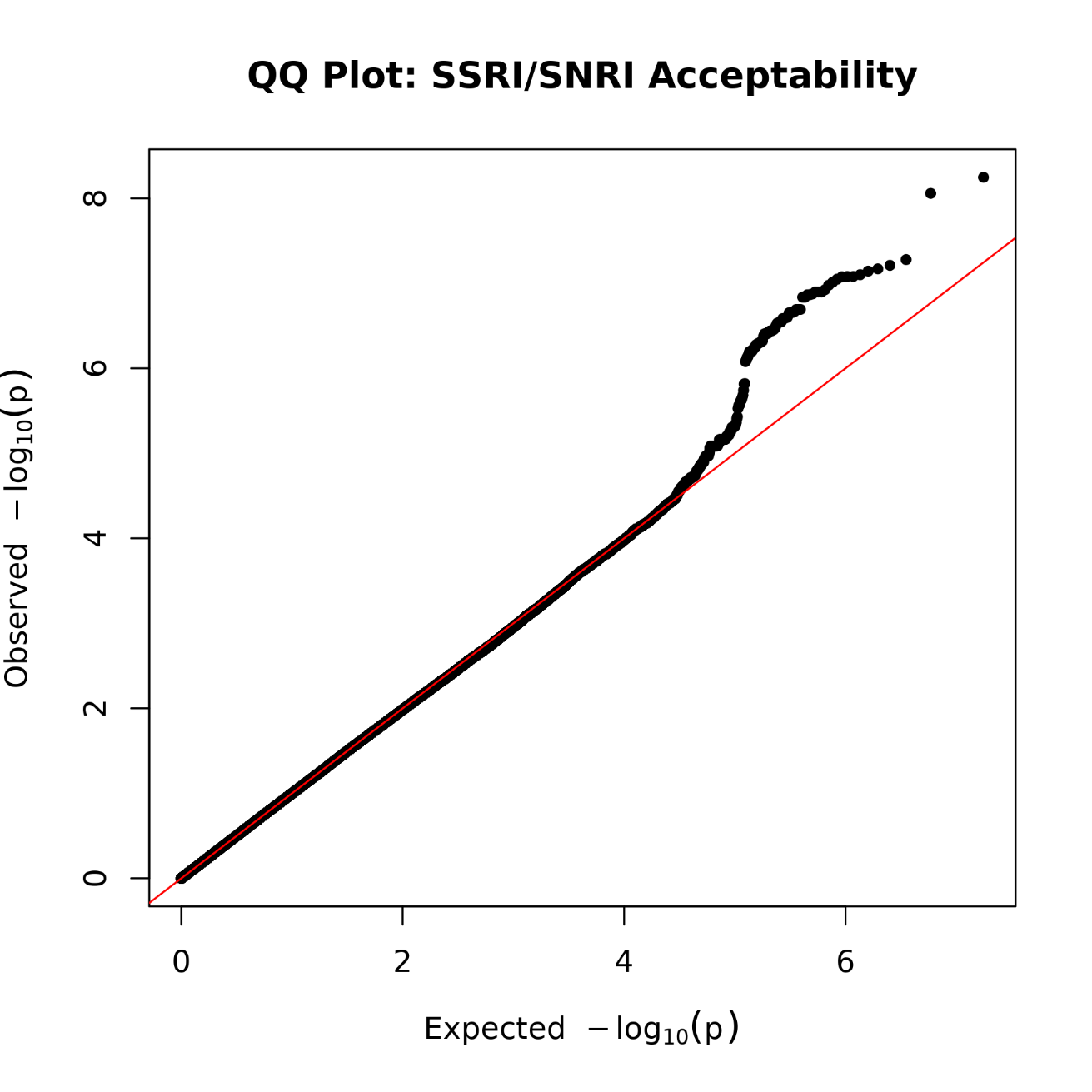

**Supplementary Figure 14**

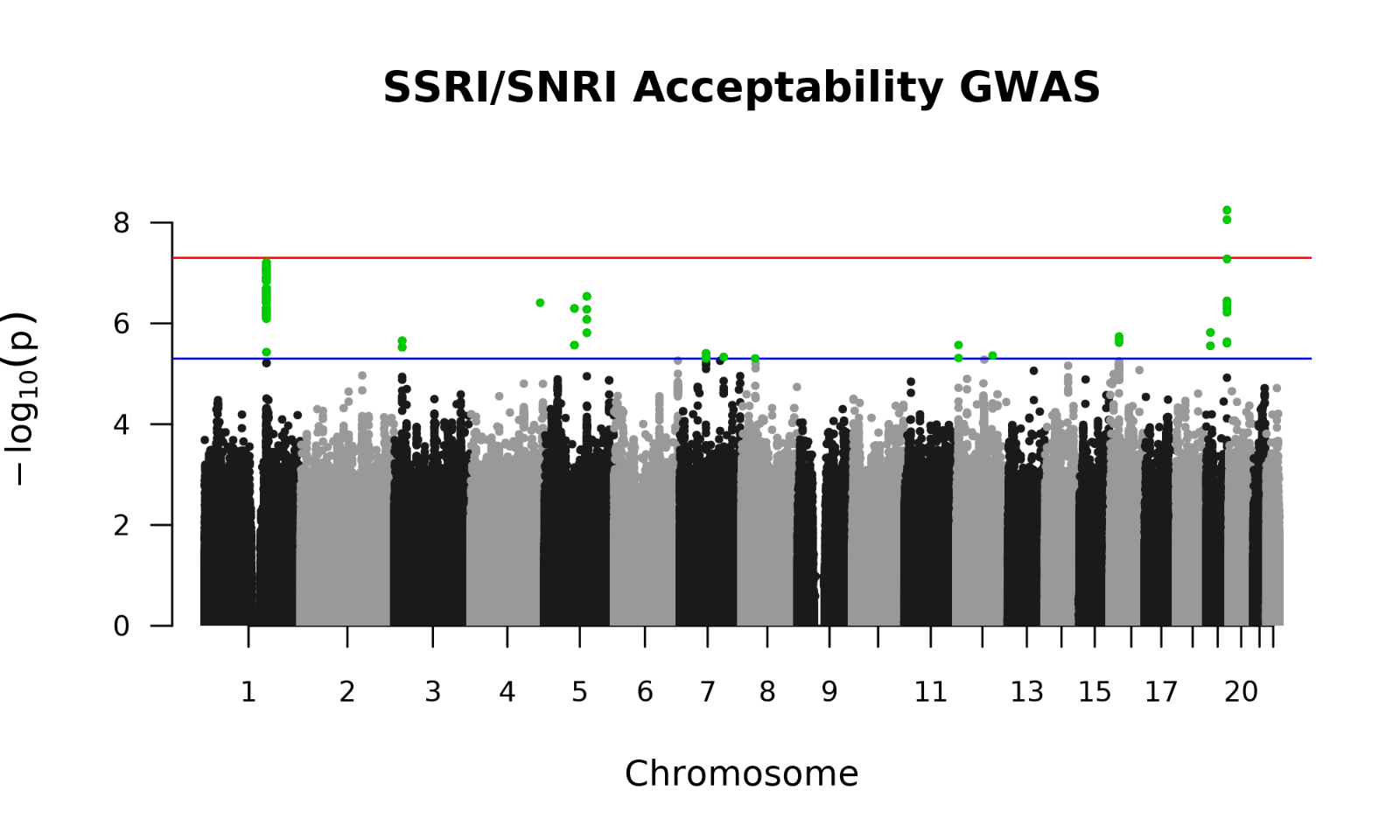

**Supplementary Figure 15 and 16. Genome-wide association study (GWAS) of SSRI acceptability.** A logistic regression GWAS was conducted comparing participants with SSRI acceptability (cases, n = 3,423) to those with non-acceptability (controls, n = 4,561). Controls were drawn from participants not classified as SSRI acceptable, excluding those with sustained SSRI use (≥360 days) or self-reported bipolar disorder. Statistical analyses were performed using the same methods as the SSRI/SNRI acceptability GWAS. Supplementary Figure 15: Quantile-quantile (QQ) plot and the genomic inflation factor (λ = 0.999) indicating minimal inflation. Supplementary Figure 16: Manhattan plot of all common SNPs. SNPs surpassing suggestive significance (p < 5.0 × 10⁻⁶) are highlighted in green. The SNP-based heritability was estimated at *h²* = 0.12 ± 0.123 (mean ± SD), based on an observed phenotypic variance of 0.25 for the binary SSRI acceptability trait.

**Supplementary Figure 15**

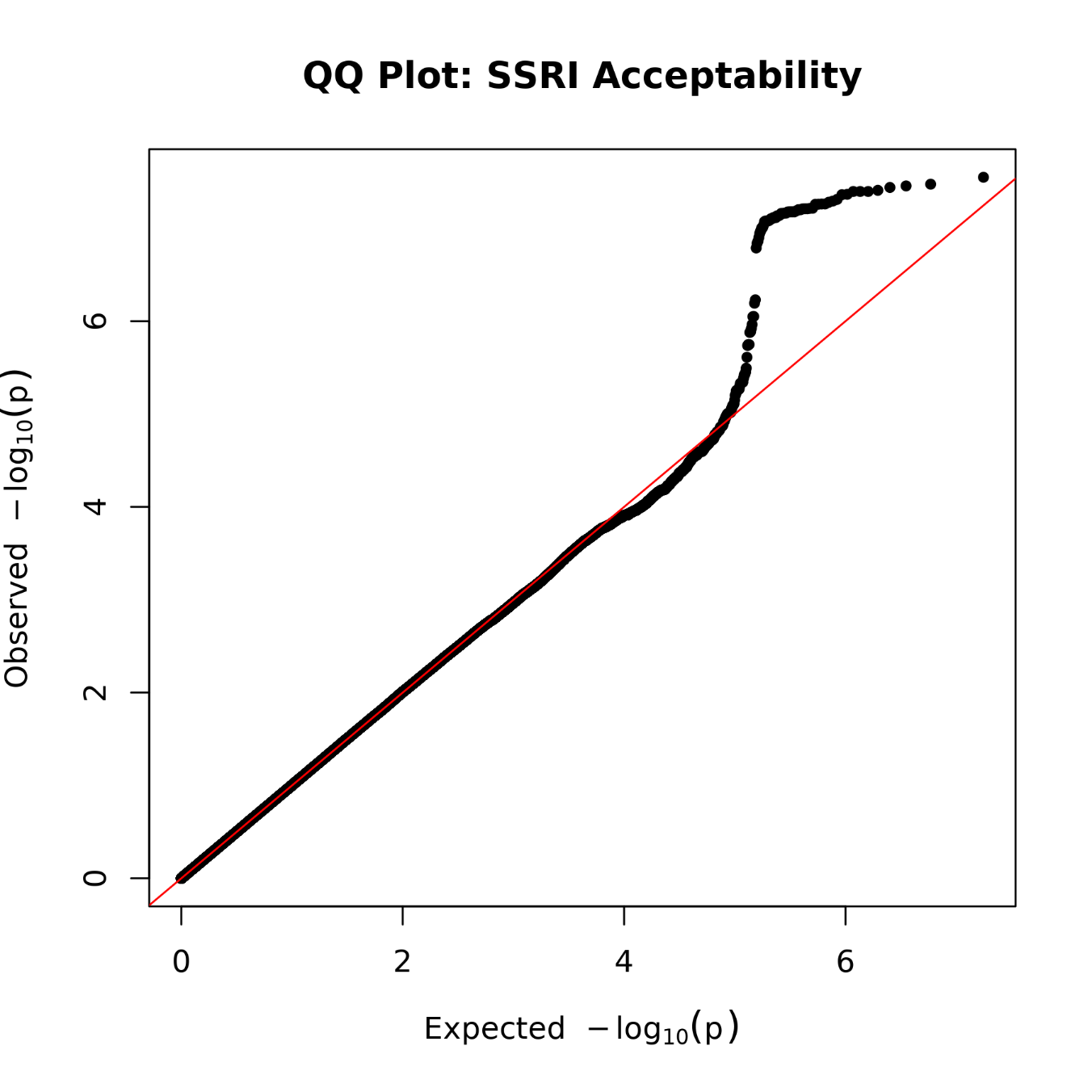

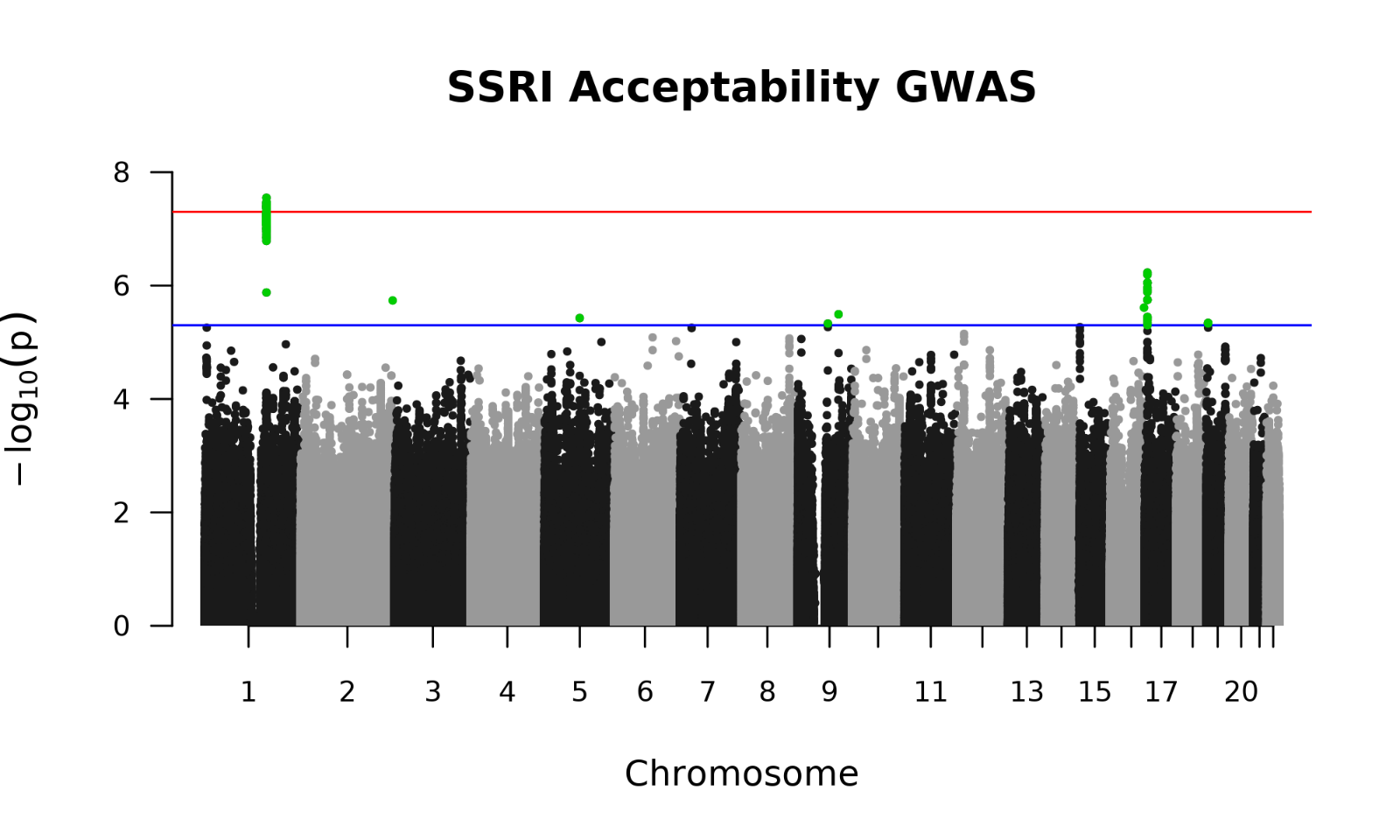
**Supplementary Figure 16**

**Supplementary Figures 17 and 18. Genome-wide association studies of self-reported antidepressant efficacy.** Supplementary Figure 17: Results from a logistic regression GWAS comparing participants who reported efficacy with either an SSRI or SNRI (cases, *n* = 8,548) versus those who reported no efficacy (controls, *n* = 935). SNP-based heritability was estimated as *h²* = 0.053 ± 0.098 (posterior mean ± SD), based on an observed phenotypic variance of 0.08 for the binary SSRI/SNRI efficacy trait. The GWAS showed minimal genomic inflation (λ = 0.999), and the Manhattan plot highlights SNP-level associations across the genome. Supplementary Figure 18: Results from a logistic regression GWAS focused on SSRI self-reported efficacy, comparing cases (*n* = 6,739) to non-efficacy controls (*n* = 2,744). SNP-based heritability was estimated as *h²* = 0.057 ± 0.086 (posterior mean ± SD), based on an observed phenotypic variance of 0.21. Genomic inflation was minimal (λ = 1.005), and SNPs are visualized in a Manhattan plot.

**Supplementary Figure 17**

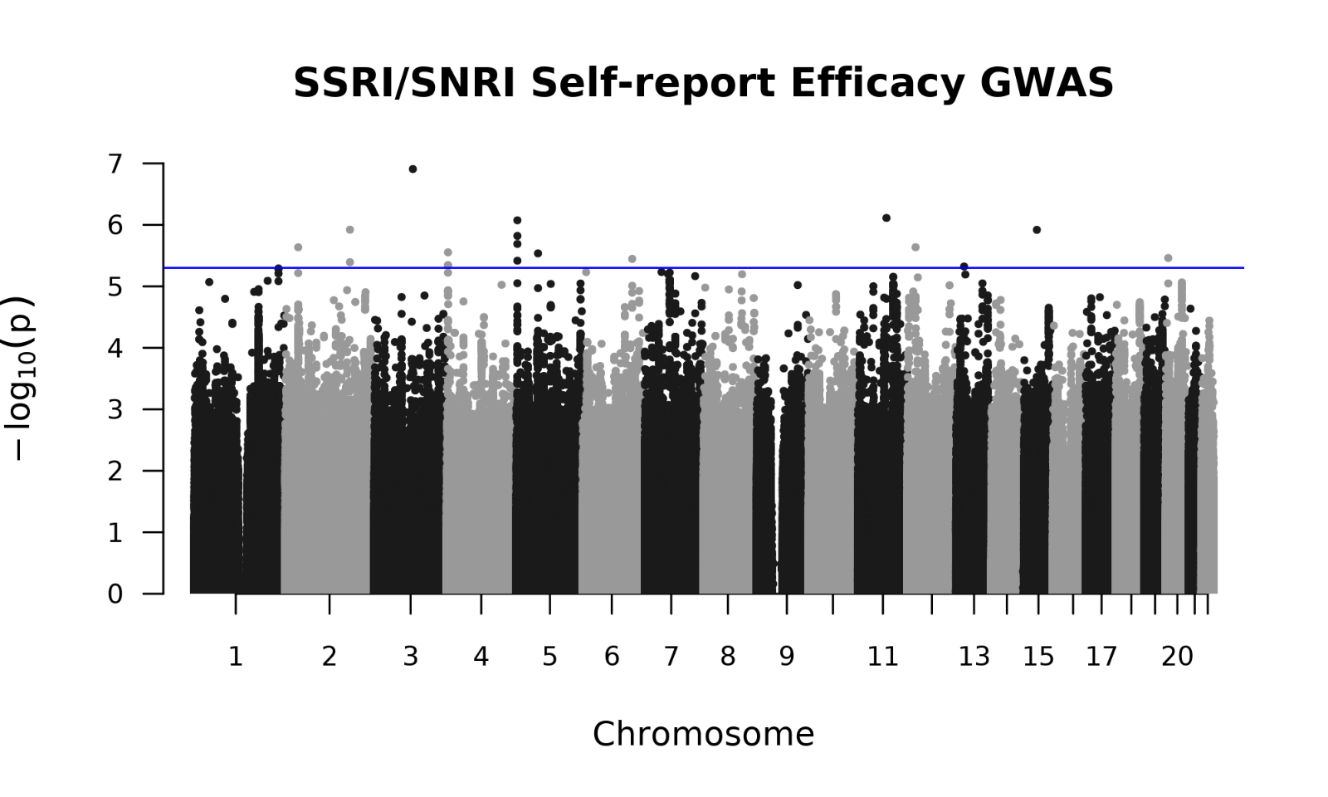

**Supplementary Figure 18**

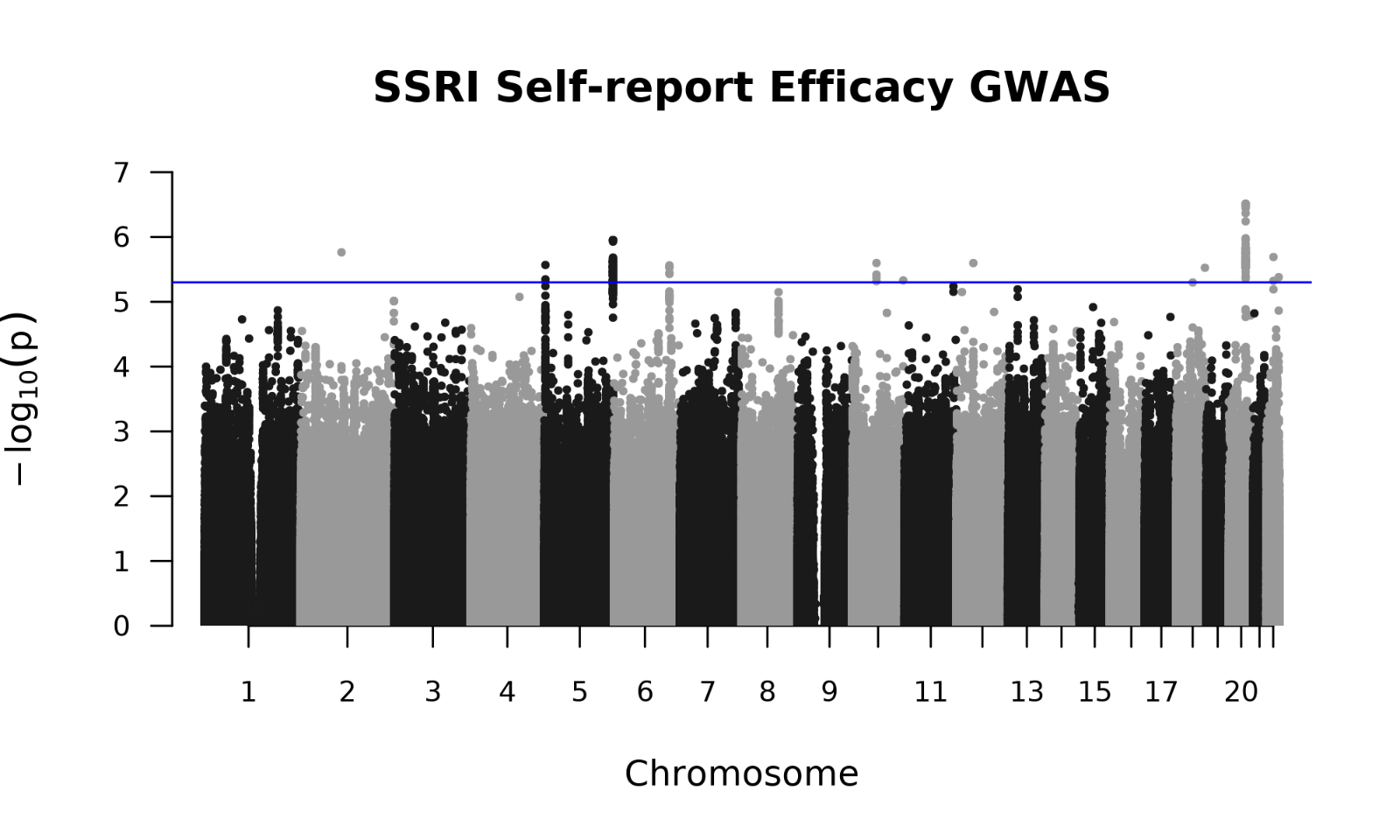
